# Dynamic ctDNA mutational complexity in melanoma patients receiving immunotherapy

**DOI:** 10.1101/2022.09.19.22280131

**Authors:** Sandra Fitzgerald, Cherie Blenkiron, Rosalie Stephens, Jon A Mathy, Tiffany Somers-Edgar, Gill Rolfe, Richard Martin, Christopher Jackson, Michael Eccles, Tamsin Robb, Euan Rodger, Ben Lawrence, Parry Guilford, Annette Lasham, Cristin Print

## Abstract

Circulating tumour DNA (ctDNA) analysis promises to improve the care of people with cancer, address health inequities and guide translational research. This observational cohort study used ctDNA to follow 29 New Zealand (NZ) unresectable advanced-stage cutaneous melanoma patients through multiple cycles of immunotherapy, to identify the breadth and complexity of tumour genomic information that ctDNA analysis can reliably report. During the course of treatment, a high level of dynamic mutational complexity was identified in blood plasma of these patients, including: multiple *BRAF* mutations in the same patient, clinically-relevant *BRAF* mutations emerging through therapy, and co-occurring sub-clonal *BRAF* and *NRAS* mutations. The technical validity of this ctDNA analysis was supported by high sample analysis-reanalysis concordance as well as by concordance between three ctDNA measurement technologies: droplet digital polymerase chain reaction (ddPCR), a custom melanoma-specific amplicon next-generation sequencing (NGS) panel and mass spectrometry. In addition, we observed >90% concordance in the detection of ctDNA when using cell-stabilising collection tubes followed by 7-day delayed processing, compared to standard EDTA blood collection protocols with rapid processing. We also found that undetectability of ctDNA at a proportion of treatment cycles was associated with both clinical benefit (best RECIST response) and prognosis (disease-specific survival). In summary, we found that multiple ctDNA processing and analysis methods consistently identified complex longitudinal patterns of clinically-relevant mutations, adding support for expanded implementation of this technology to guide in-treatment tailored cancer therapy.

## Introduction

Melanoma is known to be genomically complex [1] and to progress through complex cooperative genomic mechanisms [2], with a relatively large number of mutations relative to other tumour types [3], especially in melanoma tumours with a sun exposure aetiology. In some patients, these mutations can be detected in the blood as circulating tumour DNA (ctDNA), which can provide a convenient window into the evolution of tumour mutations over time [4]. Plasma ctDNA levels are claimed to have prognostic utility in melanoma, including a significant association between the baseline detection of ctDNA in the pre-treatment plasma sample and lower progression-free patient survival [5] and responses to both targeted therapy and immunotherapy [6-8]. Plasma ctDNA analysis can identify tumour sub-clonality and mutations associated with drug resistance [9]. Some studies have suggested that plasma ctDNA levels can reflect therapeutic response or progression in real time [6, 10-13], even prior to radiological imaging [11, 14-19]. However, a recent prospective study found that elevated ctDNA levels indicated progression earlier than imaging in only 2 out of 16 patients [12]. Nevertheless, ctDNA analysis appears, at the very least, to be emerging as a useful adjunct to clinical imaging [9, 15].

Several ctDNA technologies have been developed to track tumour mutations in plasma. Two of the most commonly utilised are droplet digital (ddPCR) [20-24] and next-generation sequencing (NGS) [25-28]. ddPCR is highly sensitive, specific and cost-effective, but only allows for a small number of defined mutations to be investigated simultaneously. In contrast, NGS allows a considerable number of genomic regions to be interrogated concurrently and can detect both known and unknown mutations but has traditionally displayed a lower level of sensitivity than ddPCR and is more expensive. A third methodology, MALDI-TOF mass spectrometry, can identify moderate numbers of specific mutations with similar sensitivity and cost to NGS [29-31]. Using these methodologies, it has been reported that the concentration of released ctDNA positively correlates with tumour size [32]. Assay sensitivity can however be impacted by delays to sample processing [33, 34] for both NGS [35] and ddPCR [36, 37]. Cell-stabilising blood collection tubes allow delayed analysis of ctDNA collected from patients living in geographically remote locations without loss of sensitivity [33].

In New Zealand (NZ) where this study is based, people from geographically-remote regions [38] as well as indigenous Māori [39, 40] and Pacific peoples [41], suffer significant inequities in cancer care [42]. In 2020, people in NZ also suffered the highest age-adjusted mortality from metastatic melanoma in the world (5 per 100 000 person-years) [43, 44]. Although the incidence of melanoma in NZ is lower in Māori and Pacific Peoples than in European New Zealanders, Māori and Pacific Peoples on average experience poorer outcomes once diagnosed [45]. This is in keeping with patterns reported among Indigenous people in other parts of the world [46, 47]. The potential to either reduce or perpetuate these cancer inequities in NZ is important to consider when assessing local use of new technologies such as ctDNA.

In this observational cohort study, we set out to identify the breadth and complexity of tumour genomic information that ctDNA analysis can reliably report. This included analysis of the consistency of information generated by different ctDNA analysis technologies (ddPCR, NGS and MALDI-TOF Mass Spectrometry), factors that may contribute to variability in mutation detection, and the effect on assay sensitivity of altering blood collection protocols to suit patients living in geographically-remote locations.

## Materials and methods

### Ethical approval, study design, patient recruitment and clinical follow-up

Twenty-nine patients with stage IV metastatic cutaneous melanoma commencing pembrolizumab or nivolumab treatment in Auckland NZ, were enrolled into the study between October 2017 and December 2019 under the NZ Health and Disability Ethics Committee ethical approval 16/NTA/180. A further 16 stage III/IV melanoma patients that underwent full surgical resection were recruited as above, but data for only two were included in this manuscript. Study numbers were dictated by the number of patients who could be recruited during the study time period. Eligible patients were either Stage III melanoma surgical patients, or stage IV melanoma patients who met the funded immunotherapy treatment criteria with radiologically measurable disease. All donors were at least 18 years of age and provided written informed consent. Patients with an unknown primary melanoma were included but patients who had been diagnosed with a second malignancy in the previous 5 years were excluded. Clinical characteristics of the stage IV melanoma patients undergoing immunotherapy are shown in Supplementary Table 1. CT imaging was conducted at approximately three-monthly intervals in keeping with local clinical practice. Clinical outcomes were analysed retrospectively and included duration of treatment and overall survival. Using RECIST version 1.1 criteria [48], patients were classified according to best tumour response achieved during the course of therapy, and their disease status at the time of final blood sample and overall survival.

### Sample collection and processing

Peripheral venous blood samples were collected from pre-surgical and subsequent clinical appointments for each surgical patient, and prior to each cycle of pembrolizumab/nivolumab therapy (cycles 1-5) and then three-monthly thereafter for immunotherapy patients. Bloods were collected into either: (i) three 10mL K_2_-EDTA vacutainer tubes, with plasma prepared within 4 hours of venesection, (ii) three 8mL Cell-Free DNA collection tubes (Roche) left at room temperature for seven days before plasma preparation. In addition, one 10mL clot activator tube was collected for each time point and processed within 4 hours of venesection for serum. To isolate plasma and serum, blood tubes were centrifuged at 1,500 x g for 10 mins at room temperature. For plasma, supernatant from all three K_2_-EDTA or Cell-Free DNA tubes were pooled, re-centrifuged at 4,000 x g for 10 mins and the supernatant plasma was stored at -80°C in 2mL aliquots. 0.25mL serum was used for Lactate Dehydrogenase (LDH) analysis conducted by the Auckland City Hospital LabPLUS clinical laboratory.

### Preparation of germline genomic DNA (gDNA) and cell-free DNA (cfDNA)

cfDNA was isolated from 5mL plasma using the QIAamp Circulating Nucleic Acid kit (Qiagen), as per the manufacturer’s protocol. Leucocyte buffy coats collected after the 1,500 x g K_2_-EDTA vacutainer centrifugation were used to prepare germline genomic DNA (gDNA) using the QIAamp DNA Blood Mini Kit (Qiagen) as per manufacturer’s instructions. Fresh frozen tumour tissue samples were available from four patients, from which tumour gDNA was isolated using a NucleoSpin Tissue Kit (Machery Nagel). Both gDNA and cfDNA were eluted in ultrapure DNase/RNase-free water (Invitrogen). DNA concentration was determined using a Qubit Fluorometer (dsDNA HS or dsDNA BR Assay Kits, ThermoFisher Scientific, MA, USA).

### NGS analysis

NGS analysis was performed to identify gene mutations present in the cfDNA isolated from plasma, using an Ion AmpliSeq HD custom melanoma panel that contained 115 amplicons, covering regions of 41 genes commonly mutated in melanoma (Supplementary Table 2). Two pools with up to 10ng of cfDNA or tumour gDNA in each pool was used to generate sequencing libraries. Amplicon primers containing unique molecular identifiers (UMTs) were attached to each DNA molecule as the sample underwent an initial three cycles of PCR, any remaining UMTs were enzymatically digested, and a universal PCR was conducted to generate the completed sequencing library. Libraries were quantified using an Ion Library TaqMan Quantitation Kit (ThermoFisher Scientific) with an HS-D1000 TapeStation (Agilent, CA, USA). Libraries were templated on an Ion Chef Instrument before three cfDNA libraries, or up to nine gDNA libraries, were pooled to a final concentration of 15-20pM and sequenced using an Ion 318 v2 semiconductor sequencing chip on a Personal Genome Machine (ThermoFisher Scientific) using standard protocols. Data was processed using Ion Reporter software version 5.18 (ThermoFisher Scientific). Each mutation was reported as variant molecular tags (VMTs), and the mutational frequency defined as variant allele fraction (VAF) where the VMTs were calculated as a proportion of all unique molecular tags (UMTs). Following NGS analysis, all variants were visually verified at the level of individual reads in BAM files using the Integrated Genome Viewer [49].

### MALDI-TOF Mass Spectrometry analysis

For a subset of samples, MassARRAY UltraSEEK MALDI-TOF Mass Spectrometry analysis was performed to further orthogonally validate the detection of specific ctDNAs. Up to 10ng of cfDNA sample was analysed with the UltraSEEK MassARRAY melanoma V2 panel (Agena Biosciences [50]) following the manufacturer’s instructions. Data analysis was performed using Typer software version 4.0.26.74 (Agena Biosciences).

### ddPCR Analysis

Custom ddPCR assays were performed to orthogonally validate and quantify mutations identified by NGS using six custom ddPCR assays designed to detect DNA mutations encoding: *BRAF* V600E, *BRAF* V600K, *BRAF* K601E, *NRAS* Q61K, *NRAS* Q61R and *KIT* L576P (Supplementary Table 3). Additional ddPCR assays to detect *TERT* C228T and *TERT* C250T were used as previously described in [51]. ddPCR reactions were carried out with a Bio-Rad QX200 system, in a 20μL reaction with 1xddPCR Supermix for Probes (no dUTP) (Bio-Rad Laboratories), 8μL cfDNA template, 209nM HEX (wild-type) probe and 209nM FAM (mutant) probe, 626nM each of 10μM target specific forward and reverse primers (Integrated DNA Technologies). Annealing temperature optimisation was carried out for all six custom designed ddPCR assays (two examples are shown in Supplementary Fig. 1), and the following optimal amplification conditions were identified and used in subsequent samples: 1 denaturation cycle 95°C for 10 minutes, 40 cycles of 94°C for 30 seconds and 61.4°C for 1 minute (ramp rate 2°C per second) followed by a final 98°C for 10 minutes. Samples with >10,000 droplets generated were analysed using QuantaSoft version 1.0.596 software. Droplet counts are presented as the number of positive droplets per mL of input plasma and a threshold of ≥ 2.5 droplets/mL of input plasma was used for reporting the presence of ctDNA in a sample.

### Statistical analysis

All statistical analyses were undertaken using the R programming language [52]. The specific R packages were used: ggplot2 [53], survival [54], cowplot [55], survminer [56]. All hypothesis testing included in this paper used Wilcoxon rank sum tests with continuity correction.

## Results

### Longitudinal plasma ctDNA analysis

Peripheral venous blood samples were collected from 29 stage IV melanoma patients immediately before each cycle of immune checkpoint inhibitor therapy. Patients received between four and 34 cycles of pembrolizumab/nivolumab while participating in this study and each donated between four and 12 blood samples. For each blood sample, cfDNA extracted from plasma was analysed using ddPCR alongside at least one other independent genomic technology. From these analyses ctDNA mutations were identified in 28/29 (97%) of the patients (Supplementary Table 4).

### Comparison of ctDNA mutations to mutations previously detected in tumour tissue

Clinical genomic analysis had previously identified *BRAF* mutations in the tumour tissue of 11 study patients (Supplementary Table 4). Analysis of ctDNA identified the same *BRAF* mutation in the blood of all 11 patients (100% concurrence). *BRAF* ctDNA mutations were also identified in the blood of an additional 7 patients for whom clinical genomic analysis of tumour tissue had been conducted but had not identified mutations in the *BRAF* gene, and in an additional 4 patients who had no previous testing of tumour tissue. All four patients with an *NRAS* mutation and one patient with a *KIT* mutation identified in previous clinical genomic analysis of tumour tissue had the corresponding mutations identified in their plasma.

### Longitudinal monitoring of ctDNA during immunotherapy

A range of patterns of longitudinal ctDNA concentration over the course of immunotherapy were observed. These included monotonic ctDNA increases or decreases as have been described previously [10] as well as more complex patterns of change (Fig. 1). A full description of the mutations identified, and the quantitation of these over time is shown in Supplementary Fig. 2. For ten patients, a clear pattern of decreasing or increasing ctDNA concentration correlated with response to immunotherapy or with progressive disease (examples shown in Fig. 1a and 1b, respectively). For eleven patients, fluctuating ctDNA concentrations were identified, trending downwards or upwards consistent with treatment response (examples shown in Fig. 1c and d, respectively). However, two of the patients who demonstrated a downward trend of ctDNA that correlated with initial radiological assessment went on to develop intracranial metastases. For seven patients who responded to immunotherapy or had stable disease, fluctuating low levels of ctDNA (ranging between undetectable to <20 copies/mL plasma) were observed (example shown in Fig. 1e).

**Fig. 1.**
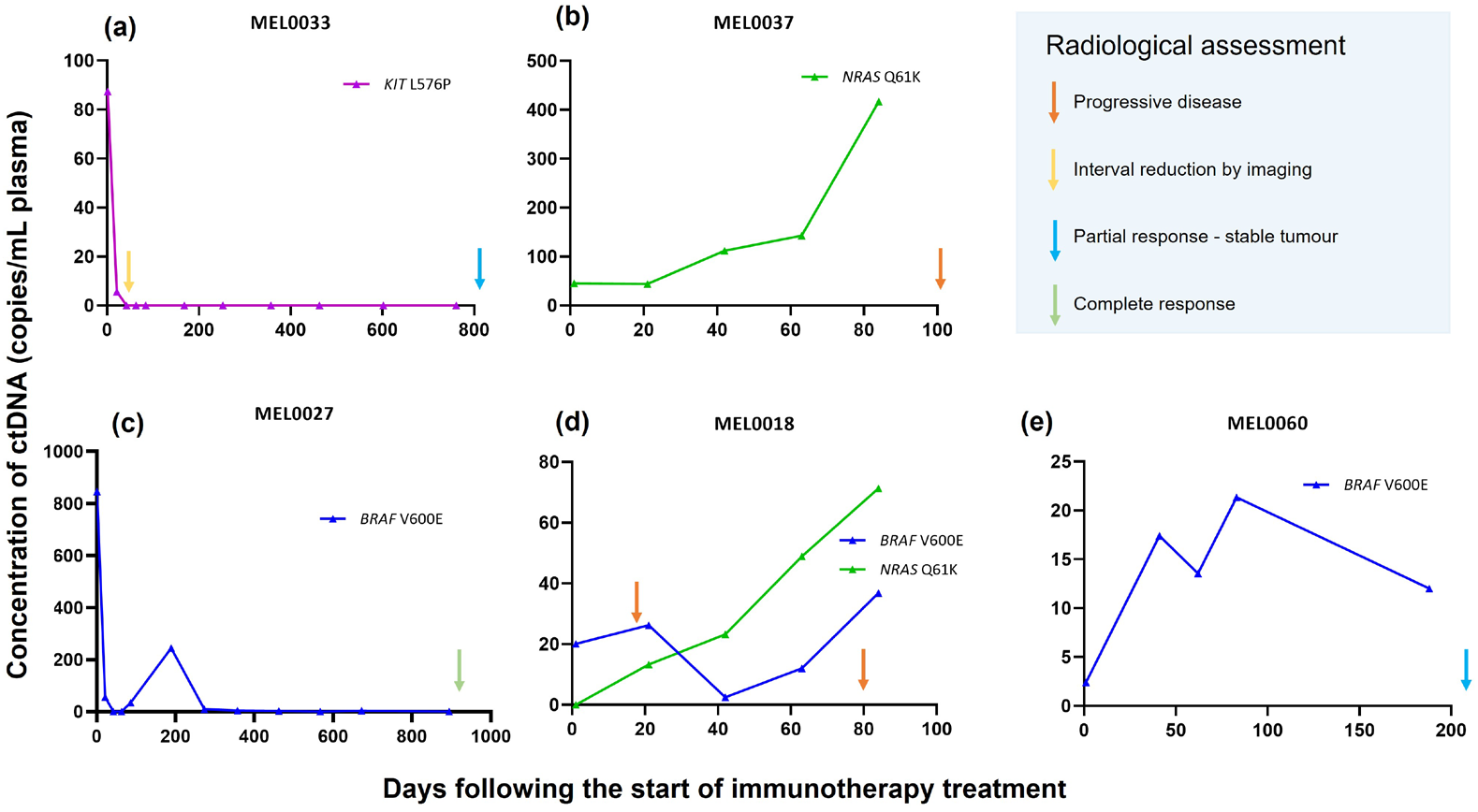
Longitudinal analysis of ctDNA levels in four melanoma patients with different responses to immunotherapy. ctDNA analysis of four melanoma patients is shown along with the CT imaging at last follow up. (a) Patient MEL0033: a decrease in ctDNA concentration was concordant with interval response to therapy confirmed radiologically, (b) Patient MEL0037: an increase in ctDNA concentration correlated with progression on therapy confirmed radiologically, (c) Patient MEL0027 demonstrated fluctuating downward ctDNA concentrations that correlated with treatment response, (d) Patient MEL0018: an initial decrease in the detectable ctDNA for *BRAF* V600E was accompanied by an increasing concentration of ctDNA for *NRAS* Q61K. (e) Patient MEL0060 continuous detection of low concentrations of ctDNA in a patient with clinically stable disease. Note the different y-axis scale in the five panels of this figure.

Since minimally invasive ctDNA analysis can be undertaken at intervals between surveillance imaging, the potential value of more frequent ctDNA monitoring assessed, as was illustrated for one patient shown in Fig. 2. Here, longitudinal analysis of *BRAF* V600E ctDNA by ddPCR was consistent with an initial response to immunotherapy followed by progression on therapy, suggested by a rise in ctDNA concentration during the period between a baseline PET-CT scan and a second PET-CT scan, which was performed following clinical indications of tumour progression (Fig. 2).

**Fig. 2.**
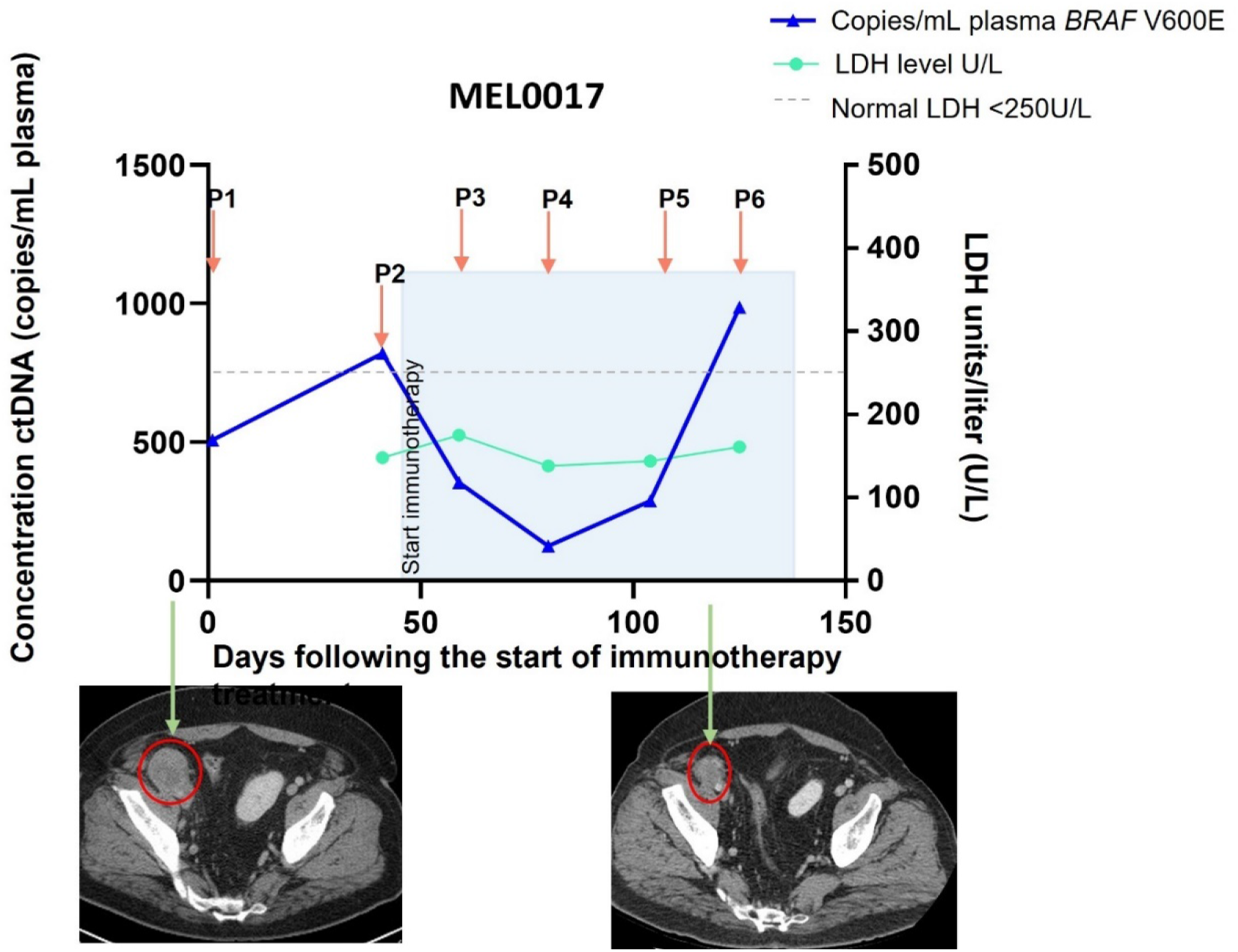
Example of a relationship between ctDNA concentration and findings from surveillance imaging: Analysis of ctDNA concentration at baseline (P1, before treatment) and the start of each immunotherapy cycle (P2-P6, orange arrows) is shown with the blue triangles and indicates an initial response (P3-P5), followed by progression on therapy occurring between the two interval PET-CT images. The serum LDH concentration (cyan line) was below the upper threshold for the normal reference range throughout (<250U/L, indicated by a dashed grey line). PET-CT images taken prior to and during immunotherapy (green arrows) suggested an interval reduction in the size of a pelvic metastatic melanoma (indicated by a red circle).

### Detection of complex tumour heterogeneity over time

Concurrent mutations in both *BRAF* and *NRAS* genes were detected in ctDNA from seven patients. These were identified by ddPCR of ctDNA from at least two plasma samples per patient collected at different timepoints. Since this high prevalence of concurrent mutations was unexpected, all ctDNA mutations detected by ddPCR were subsequently re-assessed and their presence was confirmed in the same blood plasma sample using at least one other orthogonal technology (either NGS or UltraSEEK MassARRAY). Six of the seven patients previously had genomic tumour testing performed, but mutations were only detected for two patients (one each of *NRAS* and *BRAF*). Additionally, for two of these patients, co-occurring *BRAF* and *NRAS* mutations were detected in the baseline plasma sample, but were not detected in the clinical analysis of the tumour tissue conducted two months prior to starting immunotherapy. The detection of concurrent ctDNA mutations is consistent with the presence of multiple genomic tumour cell clones within a patient, with examples shown in Supplementary Fig. 3.

In one patient (MEL0035), a *BRAF* V600E mutation was detectable in all eight plasma samples (P1-P8), with additional *BRAF* V600K and *NRAS* Q61R ctDNA mutations becoming detectable by ddPCR after the fourth immunotherapy cycle (analysis of their fifth plasma sample P5 is shown in Supplementary Fig. 4). NGS quantified the variant allele frequency (VAF) of the *BRAF* V600E, *BRAF* V600K and *NRAS* Q61R ctDNA mutations P5 as 4.6%, 6.5% and 0.2%, respectively. Given the unexpected presence of an additional *BRAF* V600K mutation potentially evolving from a *BRAF* V600E-mutant tumour, ctDNA from the P5 plasma sample was reanalysed using a third orthogonal technology, UltraSEEK MassARRAY, which also confirmed the presence of all three mutations. These observations are consistent with the *BRAF* V600K and *NRAS* Q61R mutations emerging in subclones from the original *BRAF* V600E-mutant tumour.

In three patients, no mutations were detected in either tumour tissue or ctDNA isolated from plasma taken at the time of the first cycle of immunotherapy. For one patient (MEL0022), a *BRAF* V600E ctDNA mutation only became detectable after the fourth immunotherapy cycle (P5 plasma sample), even though *NRAS* Q61R was detected from the first pre-treatment sample onwards.

### Tumour heterogeneity in stage III melanoma patients

To assess whether this mutational complexity was restricted to immunotherapy-treated patients, two stage III melanoma patients not receiving immunotherapy were also analysed. In these patients, similar degrees of ctDNA mutational heterogeneity were observed. In the first patient (MEL0026), ddPCR analysis of both a lymph node metastasis and plasma cfDNA sample identified concurrent *TERT* C228T, *BRAF* V600E and *BRAF* V600K mutations, which were confirmed by NGS analysis. The relative VAF of these three mutations were consistent with presence of a *BRAF* V600E-mutant subclone arising within a *BRAF* V600K-mutant tumour (Supplementary Fig. 5). In the second patient (MEL0011), *BRAF* V600E, V600K and K601E mutations were identified in ctDNA with the *BRAF* V600K and *BRAF* K601E becoming more prevalent 18 months after surgery. Both ddPCR and NGS analysis of cfDNA from this patient were concordant, with the NGS analysis revealing the consistent presence of variants for two independent tumour sub-clones (Supplementary Fig. 6).

### Association between ctDNA and clinical outcome

While identifying clinical associations between ctDNA level and immunotherapy response in melanoma patients was not the core purpose of this study, some interesting associations were observed. We found that those patients who were categorised as having progressive disease (PD) as a best RECIST response had higher concentrations of ctDNA in their final sample compared to those with best RECIST responses of stable disease, partial or complete response to therapy (p = 0.0003, Supplementary Figure 7 a and c). However, when analysing the *baseline* (first) pre-treatment samples, we found no significant association between ctDNA concentration and best RECIST response (Supplementary Figure 7 b). We also observed an association between disease-specific survival and ctDNA concentration in the final (Suppl. Fig 7c, p = 0.003) but not first (Suppl. Fig 7d, p = 0.65) blood samples.

Previous studies have suggested ctDNA becoming undetectable in at least one treatment time point is significantly associated with patient survival [57]. In our cohort of immunotherapy-treated stage IV melanoma patients we found that the proportion of samples with “undetectable” ctDNA (< 2.5 copies/mL plasma) was significantly lower if the patients had progressive disease than if they had stable disease, partial, or complete response to therapy (p = 0.005; Supplementary Fig. 8a). We also observed that the proportion of samples with undetectable ctDNA was significantly associated (p = 0.0009) with patients surviving 2years after commencement of immunotherapy (Suppl. Fig 8b) and that the proportion of samples with undetectable ctDNA over the course of therapy was associated with disease-specific survival (Suppl. Fig 8c, p = 0.005). For this analysis the threshold for “undetectable” ctDNA was set at 2.5 copies/mL plasma, however we found similar associations when the thresholds were set at 5, 10 or 20 copies/mL (data not shown).

We next investigated whether the relationship we had observed between patient outcome and the number of longitudinal blood samples with undetectable ctDNA could be clinically useful. We observed that undetectable ctDNA in one or more blood samples taken during the first five therapy cycles had a significant association with patient outcome (Figure 3). The possibility of using this information to guide clinical decisions at that time point is explored in the Discussion, below.

**Figure 3.**
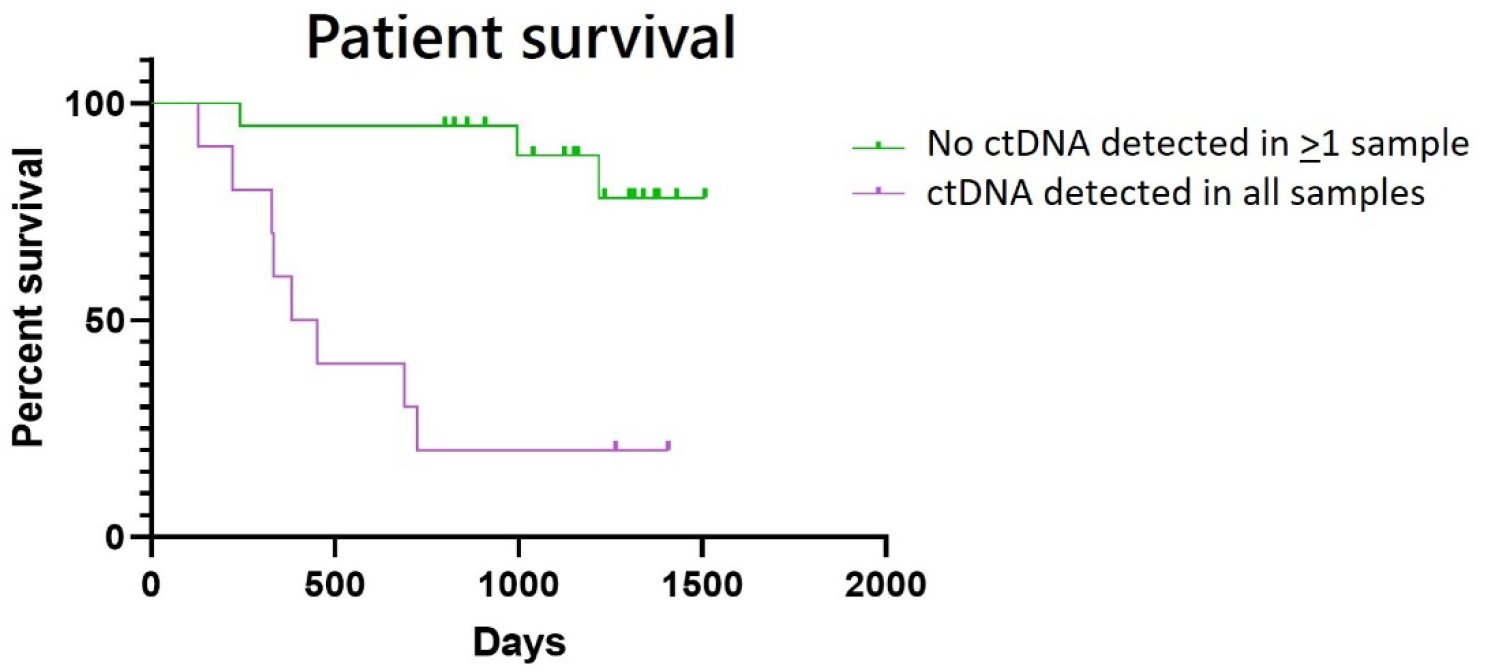
Association between undetectable ctDNA during the first five immunotherapy cycles and patient outcome. Log rank tests identified a significant positive association (P = 0.0001) between patient outcome (overall survival) and ctDNA being undetectable (< 2.5 copies/mL plasma) in one or more blood samples taken during the first five therapy cycles.

### Assessment of technical parameters of ctDNA analysis relevant to clinical implementation Consistency

Several preanalytical and analytical variables can affect ctDNA analysis [56-58]. These include time to plasma processing, cfDNA extraction protocol and platform selected for ctDNA analysis. To assess some of these factors, first, cfDNA was extracted on two different days from 38 paired aliquots of plasma, each pair of aliquots originating from the same blood sample. Although there was an overall correlation between the concentration of cfDNA isolated in the paired preparations (R^2^ = 0.96, Supplementary Fig. 9), some variability in cfDNA concentrations was observed (mean CV 13.4% range 0.3-41.3%), which is concordant with previous reports [58].

Next, we assessed how freeze-thawing of cfDNA stocks might impact the final ctDNA concentration. For 65 cfDNA samples, the ctDNA concentration was determined after one and two freeze-thaw cycles, using custom ddPCR assays. The same ctDNA was detected in the replicate ddPCRs for 62/65 (95%) of samples, and the ctDNA concentrations following one and two freeze-thaw cycles showed very good concordance (R^2^ = 0.998; Supplementary Fig. 10). For these 62 samples, the concentration of ctDNA was >4 copies/mL plasma. For the remaining 3/65 cfDNA samples, the ctDNA concentration was <4 copies/mL plasma, so the variability observed may be attributable to other factors including random sampling. On the basis of this analysis, we set the threshold for reporting the presence of ctDNA mutations in blood plasma as >2.5 copies/mL.

### Assessment of the concordance between genomic platforms for ctDNA

As was shown above, in general the ctDNA mutations detected by ddPCR assays and NGS using the AmpliSeq melanoma panel were concordant (Supplementary Table 2). Therefore, we next assessed the relative sensitivity of these two technologies in more depth for a set of mutations commonly found in melanoma. NGS analysis was performed on 42 cfDNA samples collected from 18 melanoma patients and 62 mutations were identified. For these mutations, the cfDNA sample was reanalysed using one of seven different mutation-specific ddPCR assays matching the variants detected by NGS. All 62 ctDNA mutations were identified by both technologies. The ctDNA levels quantitated by ddPCR (copies/mL plasma) and NGS (VMT, the number of individual DNA molecules identified that carry each ctDNA mutation) correlated moderately well (R^2^=0.83; Supplementary Fig. 11).

### Impact of clonal haematopoiesis on ctDNA analysis

Next, we undertook a pilot assessment of the potential impact of clonal haematopoiesis of indeterminate potential (CHIP) on the findings of this study. CHIP is the age-related clonal expansion of haematopoietic stem cells carrying mutations also found in cancer, which can be perpetuated by chemotherapy and can generate circulating DNA variants that mimic low frequency ctDNA mutations [59, 60]. CHIP can occasionally involve mutations in *BRAF* [61] and frequently involves mutations in the *TP53* gene [62]. Therefore, we analysed 20ng peripheral blood leukocytes (PBL) gDNA isolated from the first two blood collections from seven melanoma patients where ctDNA mutations in *TP53* or *BRAF* had been detected in the ctDNA by ddPCR (concentration range 2.5-4003 copies/mL plasma). For four of the seven patients, no matching mutations were detected in the PBL genomic DNA. However, the same variants were identified in PBL genomic DNA of three patients, but the DNA concentration was extremely low (range 0.091-0.370 copies/ng input gDNA), therefore it is possible that these were false positive droplets generated during ddPCR, or due to a minimal amount of ctDNA co-purified with the gDNA, as they were well below the cutoff set in this study for reporting a ctDNA mutation of >2.5 copies/mL plasma (Supplementary Table 5).

### Modification of plasma collection processes for patients in geographically remote locations

Finally, we investigated the use of cell-stabilising blood collection tubes, which are reported to allow up to 14 days delay in laboratory processing for plasma, for ctDNA analysis. This is important to assess in a NZ context, in order to implement ctDNA monitoring of patients located in geographically-remote regions. Thirty-three blood samples from 18 donors were collected at each time point in both EDTA and cell-stabilising tubes. For each patient, the EDTA tube was processed to extract plasma within four hours or after seven days for the cell-stabilising tubes. cfDNA was isolated from frozen aliquots and then ddPCR assays performed to measure the concentration of ctDNA in the samples. This identified the same ctDNA mutations in 30/33 (91%) of the patient samples collected using the cell stabiliser tubes as were detected using the standard EDTA protocol, and the concentrations showed high concordance (Supplementary Fig. 12). For three samples, ctDNA was only detected in either the EDTA tube (n=2) or the cell-stabilising tube (n=1). Since the measured ctDNA levels these samples was low (<13 copies/mL), factors other than the collection protocol may have contributed to this variability.

## Discussion

Plasma cfDNA analysis in advanced-stage melanoma patients detected complex mutational patterns over the course of immunotherapy treatment. Some of these mutations appeared to be clinically actionable. ctDNA analysis identified over 90% of the *BRAF* mutations previously found by clinical genomic testing of tumour tissue, as well as *BRAF* mutations in an additional 7 patients (24%) where prior clinical testing had not detected these mutations in the tumour. Potentially, the *BRAF* mutations detected in ctDNA but not tumour tissue may have evolved since the tumour tissue was tested or may have been present in low cellular fractions or unsampled regions of heterogeneous tumours/metastases.

Previous studies have suggested that plasma ctDNA levels reflect therapeutic responses in real time [6, 10-13]. However, our study was not designed to quantify this type of real-time temporal relationship between ctDNA concentration and treatment response, since clinical practice at the time of the study did not allow clinical imaging and venesection for ctDNA analysis to be precisely coordinated. Nevertheless, trends in ctDNA concentration for a subset of patients concorded with clear response or progression assessed according to RECIST criteria (e.g. Fig 1).

In some patients, we observed longitudinal variability in ctDNA levels. Although this could have been due to technical artefacts [63], we showed that in general our ctDNA analysis methods were technically reproducible. In particular, our results suggest that quantitation of ctDNA mutations is relatively robust to variability in cfDNA preparation efficiency, but that the number of freeze-thaw cycles may affect the detection of very low concentration ctDNAs. Based on this analysis, for the assays used here we would repeat any result of <2.5 ctDNA copies/mL plasma. CHIP could also conceivably affect measured ctDNA concentrations [59, 60], however based on our results, we believe that CHIP is unlikely to contribute significantly to longitudinal variability observed here. Therefore, we speculate that the dynamic mutational complexity seen in blood samples from some patients may reflect authentic biological changes in multiple evolving metastatic lesions with differing functional states and variable responses to treatment. Levels of detectable ctDNA are also reportedly affected by variable first pass metabolism of cfDNA [64], by progressive tumour desmoplasia blocking the export of ctDNA [64, 65], and by anatomic compartmentalisation such as the blood-brain barrier [66]. In our study, patients whose lack of response to treatment was not accompanied by a clear increasing trend in ctDNA levels had intracranial metastases (for example patient MEL0022, Supplementary Fig. 3).

Identifying clinical associations between ctDNA level and immunotherapy response in patients was not the core purpose of this study. Nevertheless, we observed that ctDNA concentration in each patient’s final blood sample of the study was significantly associated with patient outcome; which is not surprising given that for many patients the study endpoint coincided with either clear treatment response or progression on therapy. A small number of previous trials [5, 57] have found that baseline ctDNA levels are associated with patient outcome, however we did not see this association in our cohort. We also observed that the frequency of low or undetectable ctDNA in each patient’s set of longitudinal blood samples was associated with outcome, which concords with previous studies [57]. Potentially, consistent ctDNA ‘clearance’ from patients’ blood indirectly indicates the ability of an immune response to be mounted following checkpoint inhibitor treatment. Our observation that undetectable ctDNA in any blood sample taken during the first five therapy cycles had a significant association with patient outcome may support this. Potentially, this could guide escalation of infusion therapy (for example selection of combination therapy over a single agent) and/or guide escalation in imaging to monitor treatment effect (which may for example prompt switching to a salvage management option). The general clinical utility of ctDNA will be informed by current international clinical trials. The utility of ctDNA in the specific NZ clinical context described here now needs to be assessed in prospective studies.

Our NGS analysis identified substantial genomic heterogeneity in samples from individual study patients, in line with the complex mutational landscape of advanced metastatic melanoma identified in previous ctDNA studies [1, 9]. Interestingly, longitudinal ctDNA analysis in our study revealed the apparent evolution of this genomic heterogeneity over the course of treatment, which may indicate sub-clonal mutations progressively superseding one another through the selective pressure of multiple cycles of immunotherapy. The detection of multiple concurrent *BRAF* mutations in the ctDNA from patients in our study, and also of concurrent *BRAF* and *NRAS* mutations, is inconsistent with the previously accepted dogma that these mutations are mutually exclusive [1]. However, discordant *BRAF* status in multiple metastatic melanoma lesions from single patients has been reported in melanoma [1, 67], as have concurrent *BRAF* and *NRAS* mutations, where the *NRAS* mutation developed as a resistance mechanism in patients undergoing tyrosine kinase inhibitor (TKI) treatment [68, 69].

Another important finding of this study relates to the selection of ctDNA mutation(s) to assay, when the intention of ctDNA analysis is to follow treatment response. In some studies, a single oncogenic driver mutation present at an early timepoint is selected for tracking treatment response [17, 23]. However, our results suggest that for a proportion of advanced melanoma patients, the monitoring of a single target ctDNA with ddPCR may not fully represent the tumour dynamics occurring in the patient, and the use of NGS intermittently could allow the detection of any novel mutations arising.

The use of multiple orthogonal ctDNA analysis methods provides a higher level of confidence for reporting ctDNA variants detected at low levels, as suggested previously [70]. The custom Ampliseq NGS panel used in this study, which was selected due to its high reported sensitivity [71], proved only marginally less sensitive than ddPCR. We found that the results of ctDNA analysis using this NGS panel was both internally reproducible and highly concordant with ddPCR analysis. In general, previous studies of *advanced* cancer, as in our study, report a high frequency of ctDNA detection and high concordance between different analysis platforms [70]. Conversely, some studies of smaller *early-stage* tumours, which may generate lower concentrations of ctDNA have identified lower concordance between different ctDNA technologies and between different commercial test vendors [72, 73]. Despite our custom NGS panel’s apparent sensitivity, we suggest that alternative ctDNA analysis methods could further increase sensitivity in the future, such as utilising DNA fragment length differences [74], analysis of ctDNA methylation [75] and reduced amplification of wild-type DNA through blocking oligonucleotides [76]. Further improvements in sensitivity will be especially important when ctDNA analysis is used for relapse surveillance, detection of minimal residual disease or diagnostic screening.

Similar to other studies [76], we found that ctDNA detection in blood samples collected in cell stabilising tubes was possible without significant loss of test sensitivity. As these allow a delay in processing of up to 7 days, they can be applied to support ctDNA testing for geographically-remote patients [32], including some Māori [43, 44] and Pacific People [45] who suffer significant cancer outcome inequities in NZ. In addition, ctDNA analysis undertaken in the intervals between surveillance imaging may provide valuable supplementary information to guide clinical care during health service disruptions, such as those introduced during the COVID-19 pandemic.

In summary, we have shown that plasma cfDNA analysis can detect complex tumour mutational patterns and their evolution over time during treatment, a subset of which appear to be clinically informative and potentially actionable. We found that orthogonal ctDNA technologies generate largely concordant results and can allow a higher level of sensitivity by increasing confidence in ctDNA variants detected at low levels. The potential for ctDNA to be used alongside clinical imaging in a stretched health service, and the suitability of delayed processing protocols for geographically-remote patients, positions ctDNA testing as a valuable technology in our drive towards cancer care equity.

## Data Availability

All data produced in the present study are available upon reasonable request to the authors

## Acknowledgements

We thank all patients who donated samples and clinical data to our study.

We also thank Sharon Pattison (Southern District Health Board), Gareth Rivalland (Auckland City Hospital), and Richard King (Canterbury Health Laboratories) for clinical input, Rob Day (University of Otago), Stephen Wong (Peter MacCallum Cancer Centre), Elin Gray, Lesley Calapre, Mel Ziman (Edith Cowan University), and Nicola Dryden for their scientific contribution, Alice Rykers and Phillip Shepherd (Te Ira Kāwai - Auckland Regional Biobank) for coordinating patient consent and collection. This work was conducted with assistance from Grafton Clinical Genomics, Te Ira Kāwai - Auckland Regional Biobank, LabPlus, North Shore and Counties Manukau Hospital. This work was financially supported by the Cancer Research Trust NZ, Healthier Lives National Science Challenge, The Maurice Wilkins Centre, William Staunton Scholarship Fund and the University of Auckland School of Medicine Foundation.

## Supplementary Data

**Supplementary Table 1.** Clinical characteristics of melanoma patients. Showing average patient age, sex, self-reported ethnicity, tumour stage, whether clinical genomic tumour testing was performed and patient treatment.

| Clinical characteristics of stage IV malignant melanoma patients (n = 29) |  |
| --- | --- |
| Average age (years) | 66 |
| **Sex** |  |
| Male | 21 (72.4%) |
| Female | 8 (27.6%) |
| **Ethnicity** |  |
| NZ European | 26 (89.6%) |
| NZ Māori | 2 (6.9%) |
| European other | 1 (3.5%) |
| **LDH concentration (first sample) Upper Limit of normal 250U/L** |  |
| >upper limit of normal | 15 (51.7%) |
| <upper limit of normal | 4 (13.8%) |
| not performed due to haemolysis | 10 (34.5%) |
| **American Joint Committee on Cancer (AJCC) tumor stage** |  |
| III/IV surgical* | 3 (10.4%) |
| IV | 26 (89.6%) |
| **Tumour tissue testing** |  |
| Clinical tumour testing performed | 21 (72.4%) |
| Identification of *BRAF* gene mutation | 11 (52.3%) |
| Identification of other gene mutations | 4 (19%) |
| Wild-type for tested gene mutations | 6 (28.6%) |
| **Treatment** |  |
| Surgery and surveillance then commenced pembrolizumab | 3 (10.4%) |
| Pembrolizumab | 26 (89.6%) |
| Nivolumab | 1 (3%) |
| Radiotherapy during treatment | 7 (24.1%) |
| Overall survival (status at 21/2/2022) | 18 (62%) |
| *Three stage III patients demonstrated progressive disease after undergoing surgery. Pembrolizumab treatment started immediately after surgery (n=2) or 47 weeks post-surgery (n=1) |  |

**Supplementary Table 2.**
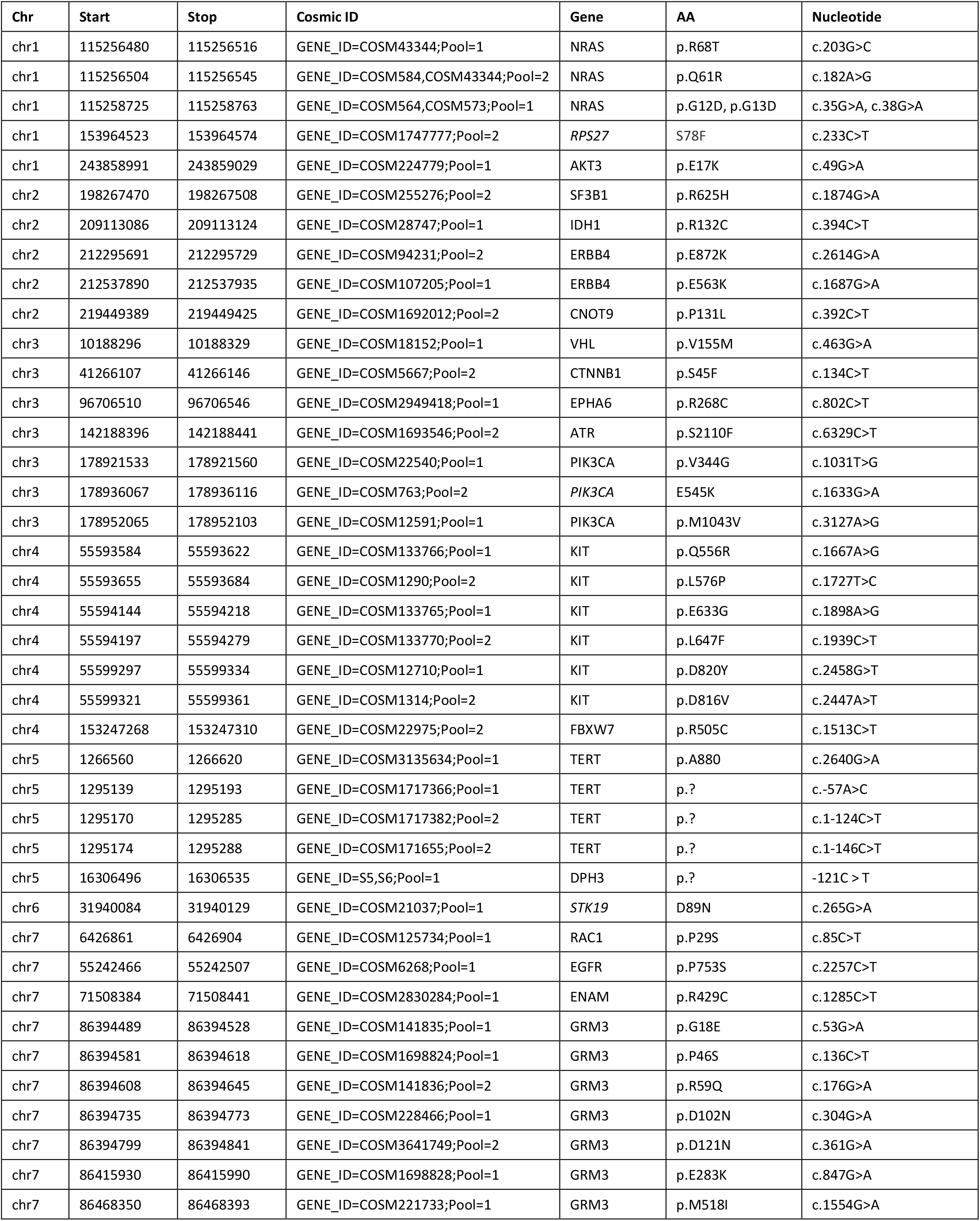
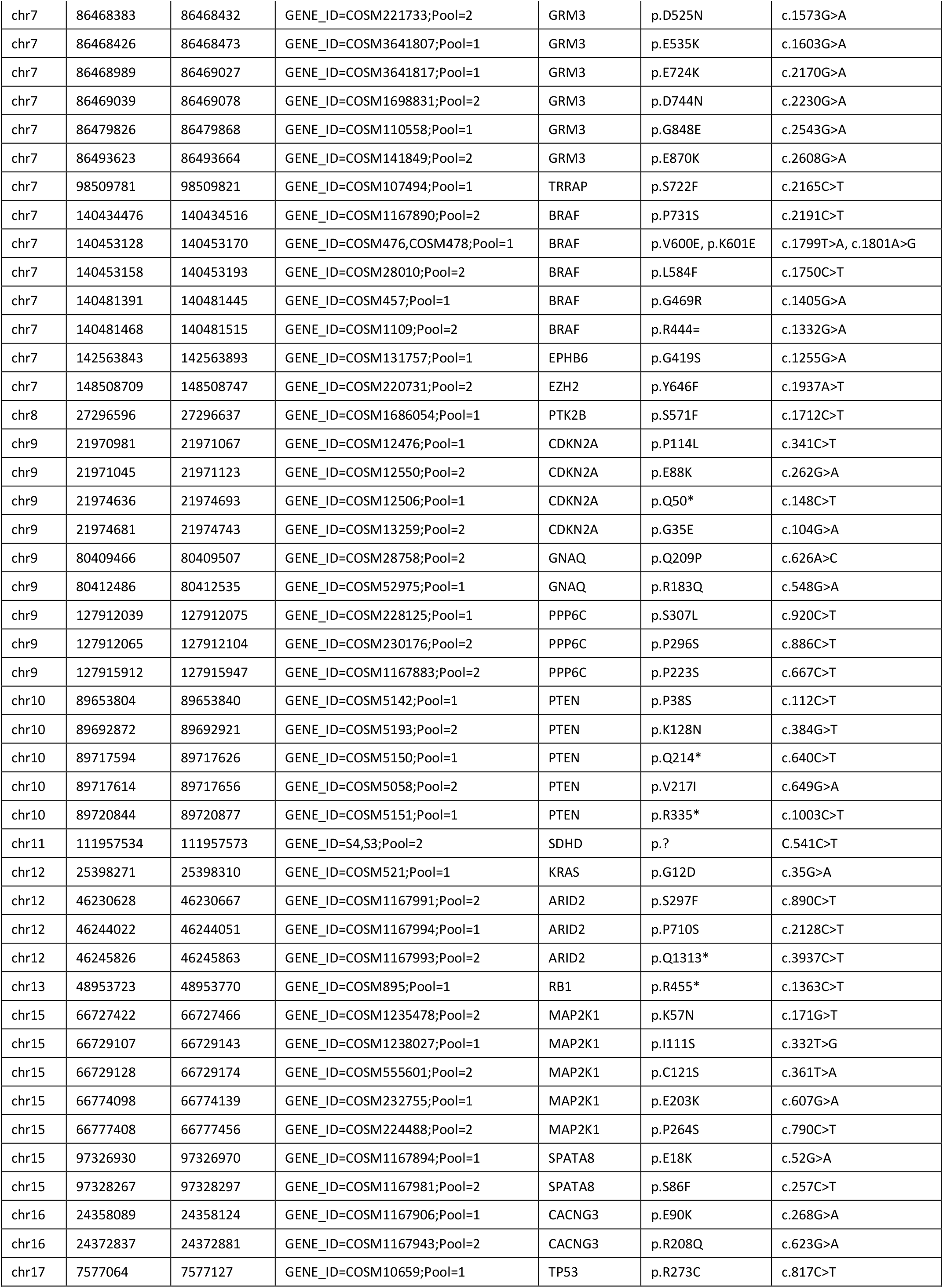
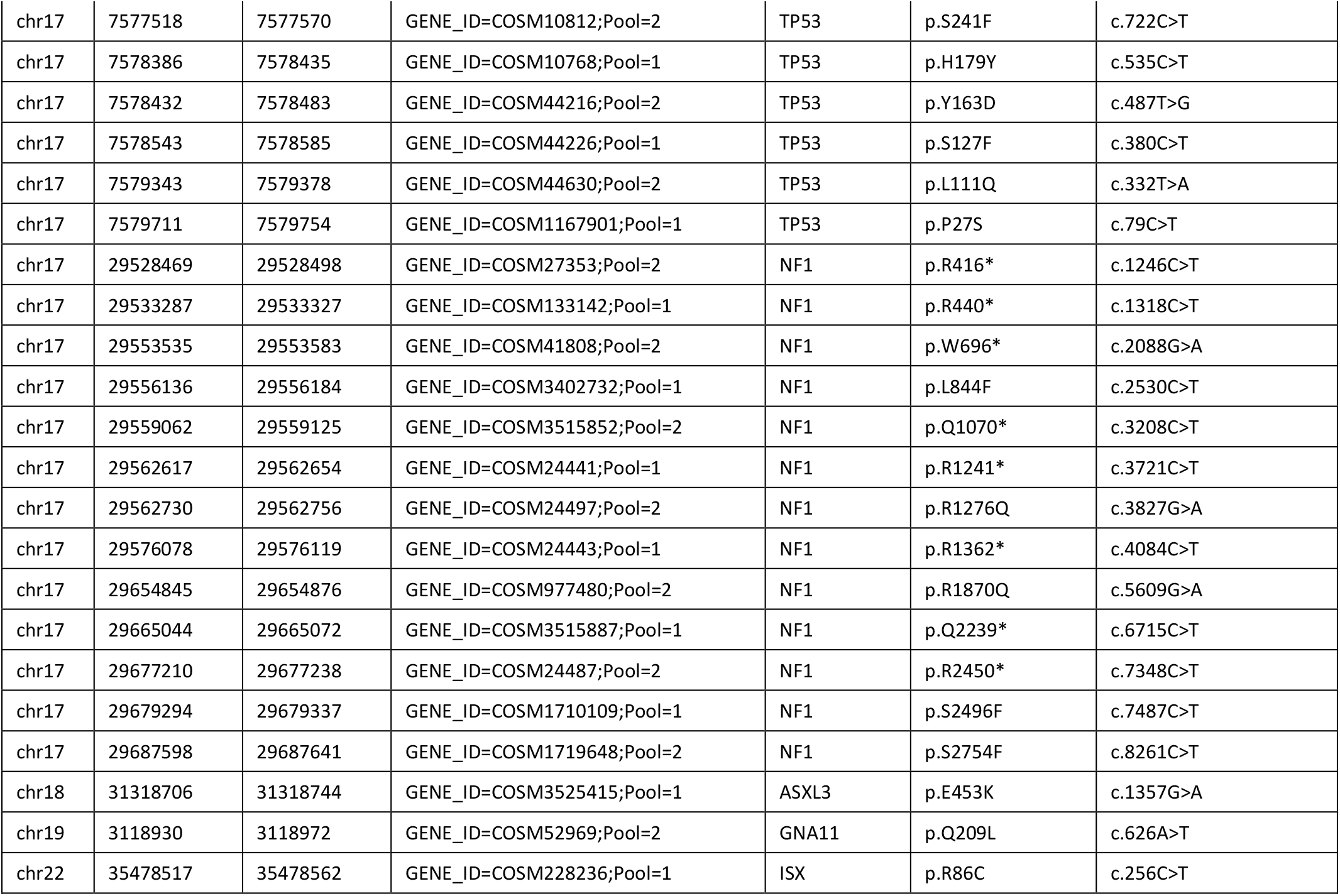
BED format description of AmpliSeq HD melanoma NGS panel (hg19 genome build).

**Supplementary Table 3.**
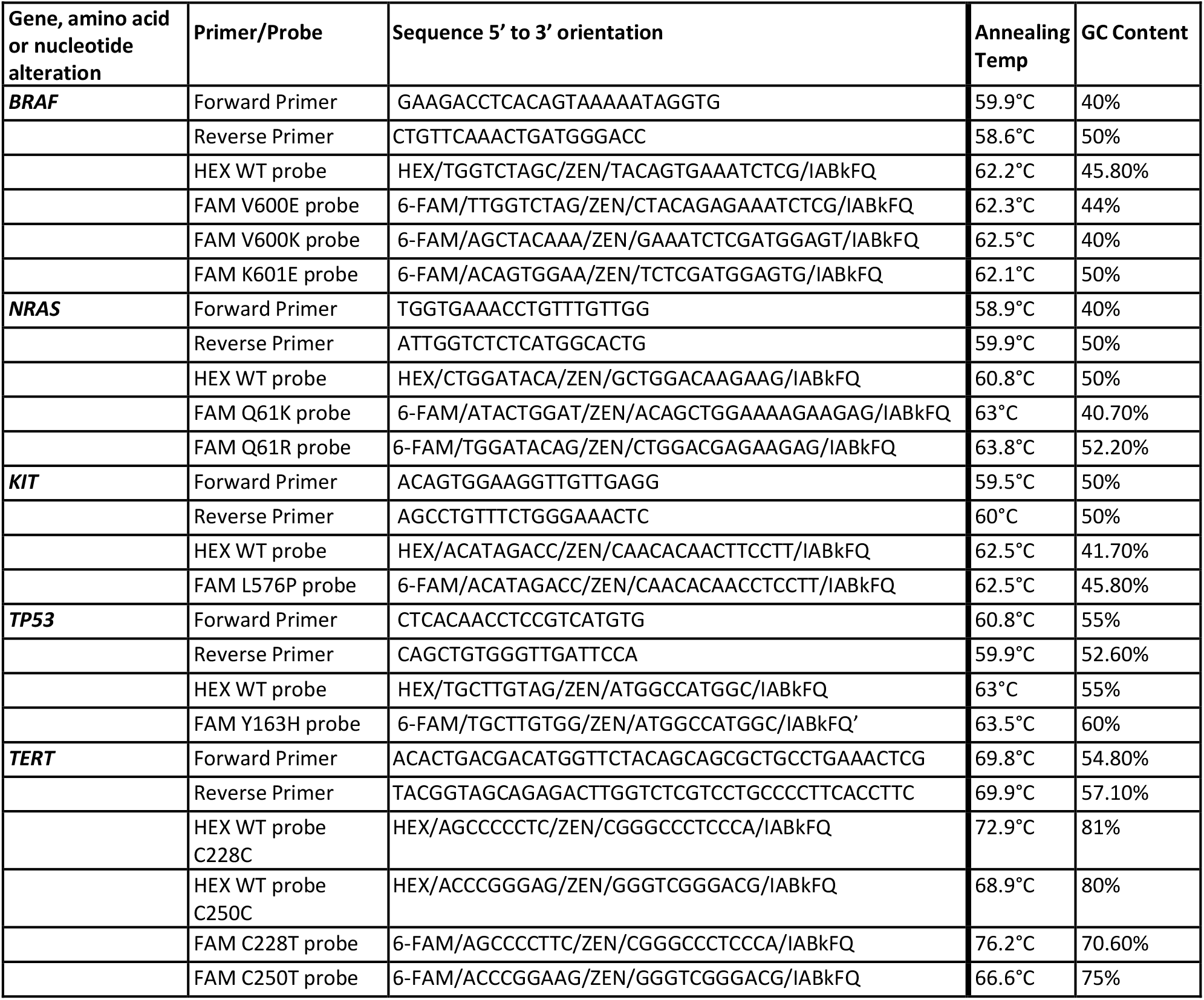
ddPCR primers and probes. Custom ddPCR assays were designed for the tracking of ctDNA in the patient’s blood. The design of these ddPCR assays followed the Bio-Rad guidelines [83]. The primer and probe sequences used for the TERT ddPCR assays were as published [58]. *=mutation leads to the formation of a stop codon. fs=frame-shift mutation. Del=deletion. Mutant bases in the FAM probes are shown in red font. FAM and HEX are the incorporated fluorophores. ZEN and IABkFQ are quenchers included in the probe design.

**Supplementary Table 4.**
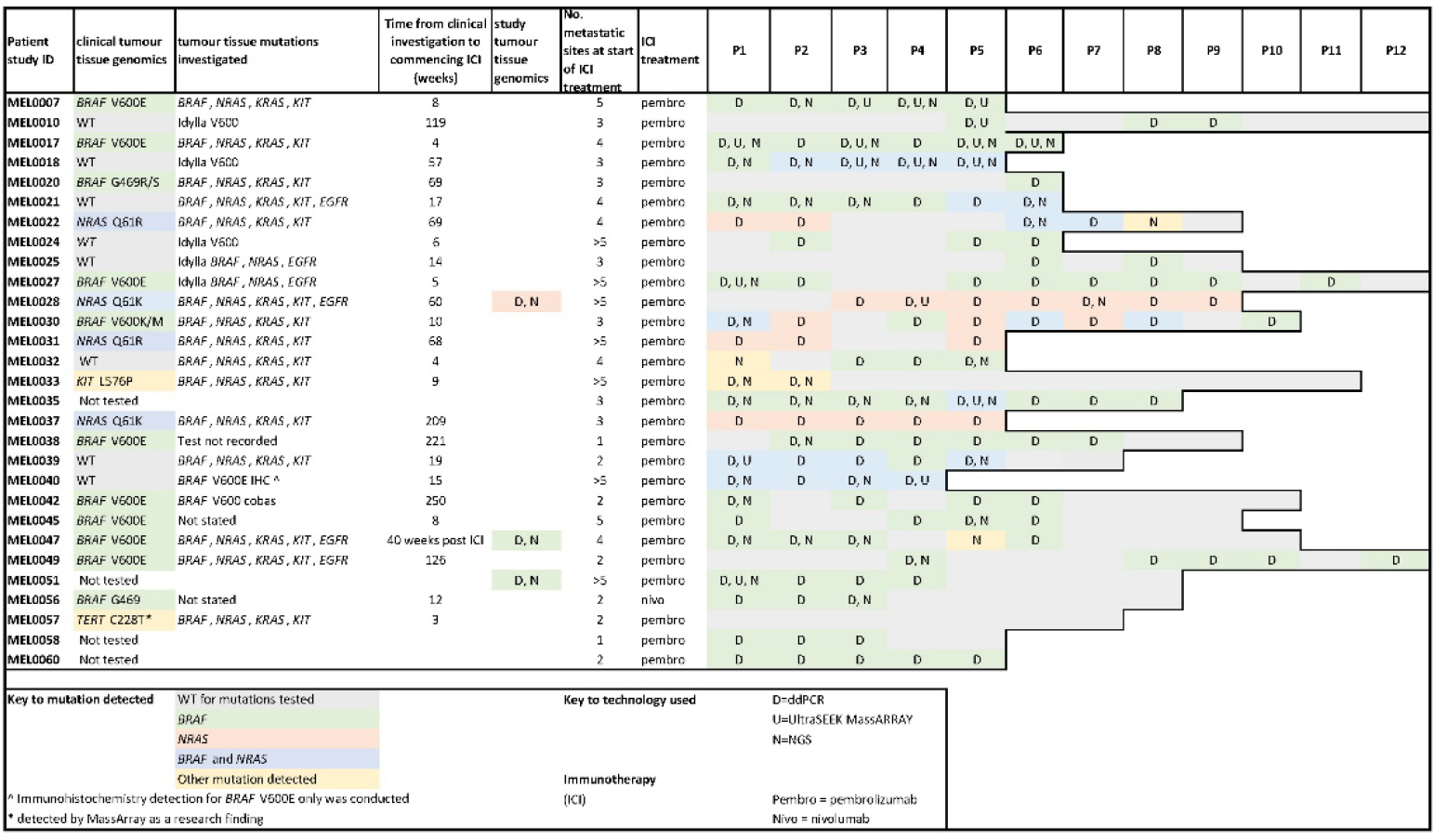
Detection of ctDNA in melanoma patient samples. For the 29 immunotherapy participants the results of clinical tumour tissue genomic analysis are shown, along with the genes investigated. For patient MEL0040, genomic analysis was performed 40 weeks after commencing immunotherapy treatment following surgical resection of an intracranial brain metastasis. For three patients, tumour tissue material was available for genomic analysis alongside the plasma samples. For these patients: MEL0028, MEL0047 and MEL0051, immunotherapy treatment was commenced 329, 54 and 48 days after tumour resection. The mutations detected and genomic platforms that were used to detect mutation are shown. The number of metastatic sites identified by radiological imaging and the immunotherapy treatment (ICI) is shown. The number of cfDNA samples collected for each patient is shown with one column per plasma sample (P#), along with the ctDNA mutations detected in each plasma sample. D, U and N indicate that mutations were detected using ddPCR, UltraSEEK MassARRAY and NGS, respectively. Coloured shading indicates the gene in which mutations were detected, according to the key at the base of the table.

**Supplementary table 5.**
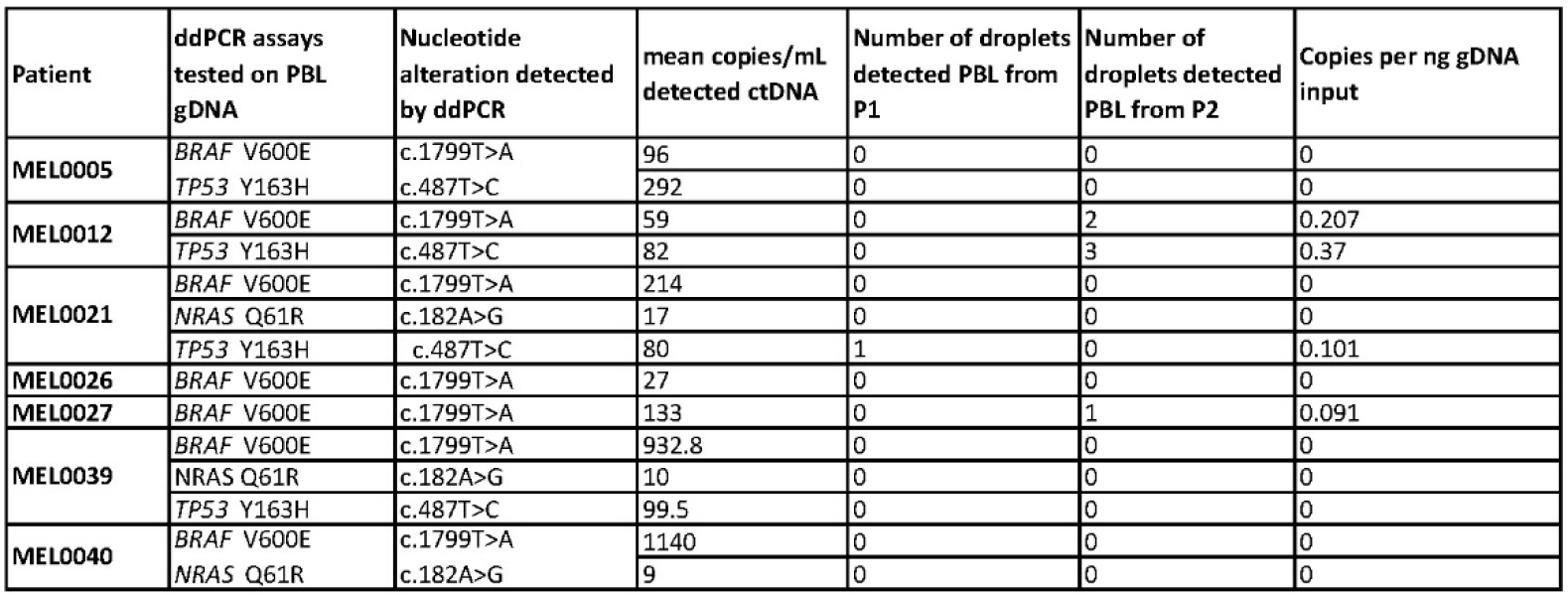
PBL gDNA analysed for mutations by ddPCR assay. PBL gDNA was used as the template for the ddPCR assay to detect mutations identified in the cfDNA. The ddPCR assay tested, the nucleotide alteration and the mean number of copies/mL plasma when the ctDNA was detected in the patient samples. The number of droplets detected in 20ng of PBL gDNA and the copies per ng gDNA input are shown. The P1 PBL sample was collected prior to commencing immunotherapy, and the P2 PBL sample was collection immediately prior to the second immunotherapy treatment.

**Supplementary Fig. 1.**
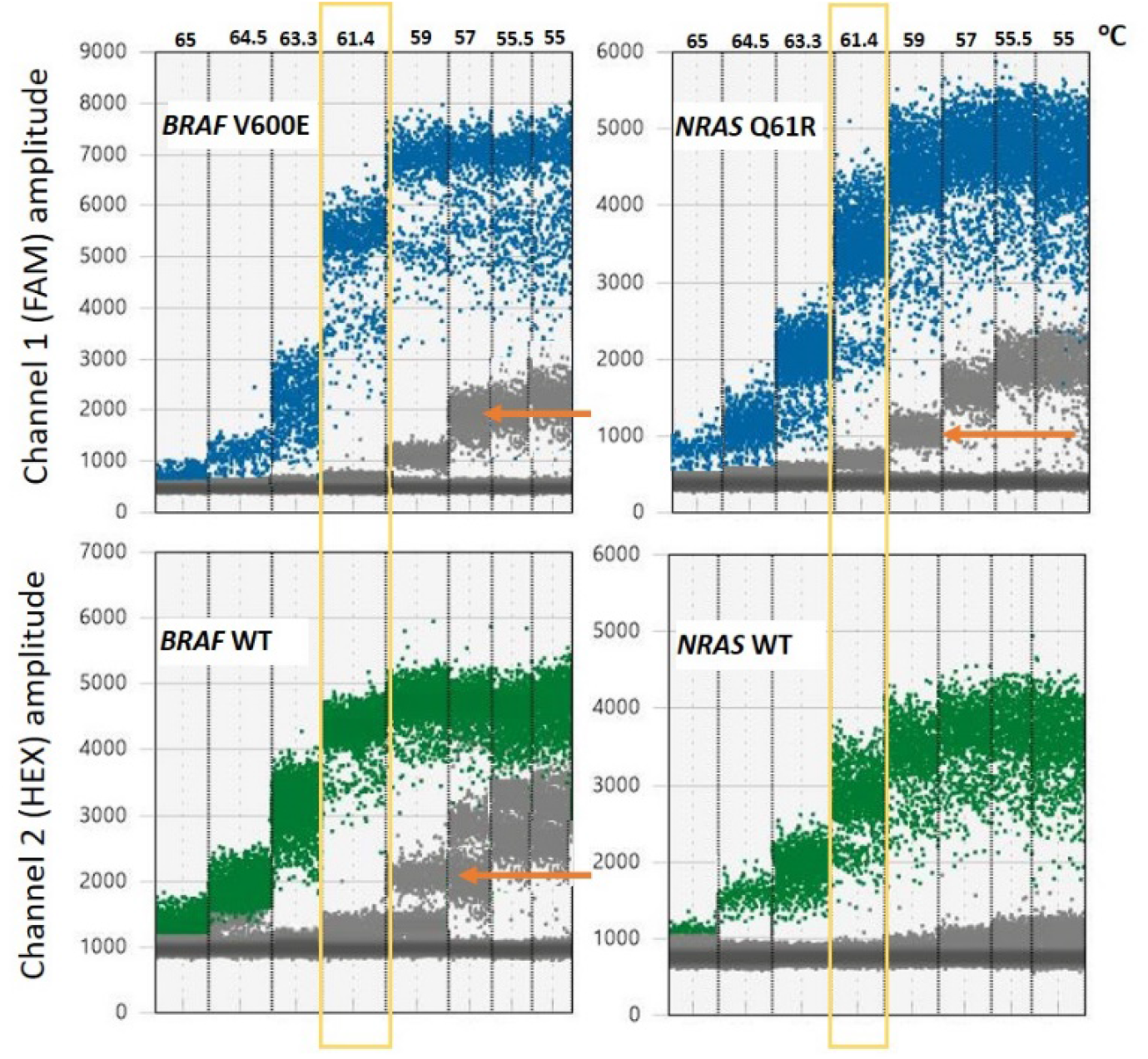
Optimisation of ddPCR rare event detection assays. Results from the optimisation of the annealing temperature for the *BRAF* V600E and *NRAS* Q61R ddPCR assays with a positive control template. Data visualised in a 1-D plot showing separation of the negative and positive droplets for *BRAF* V600E/*BRAF* WT and *NRAS* Q61R/*NRAS* WT ddPCR assays over the annealing/extension temperature gradient (55-65°C). Both fluorescent channels are shown separately. Blue droplets represent the detection of DNA with the mutant sequence. Green droplets represent the detection of DNA with the WT sequence. Grey droplets have no target DNA amplification. Optimal separation of the FAM and HEX positive droplets was achieved for both assays with the probe annealing temperature of 61.4°C, as indicated by the yellow boxes. Non-specific hybridisation by the probe is shown as additional clusters of negative (grey) droplets (examples indicated with orange arrows).

**Supplementary Fig. 2.**
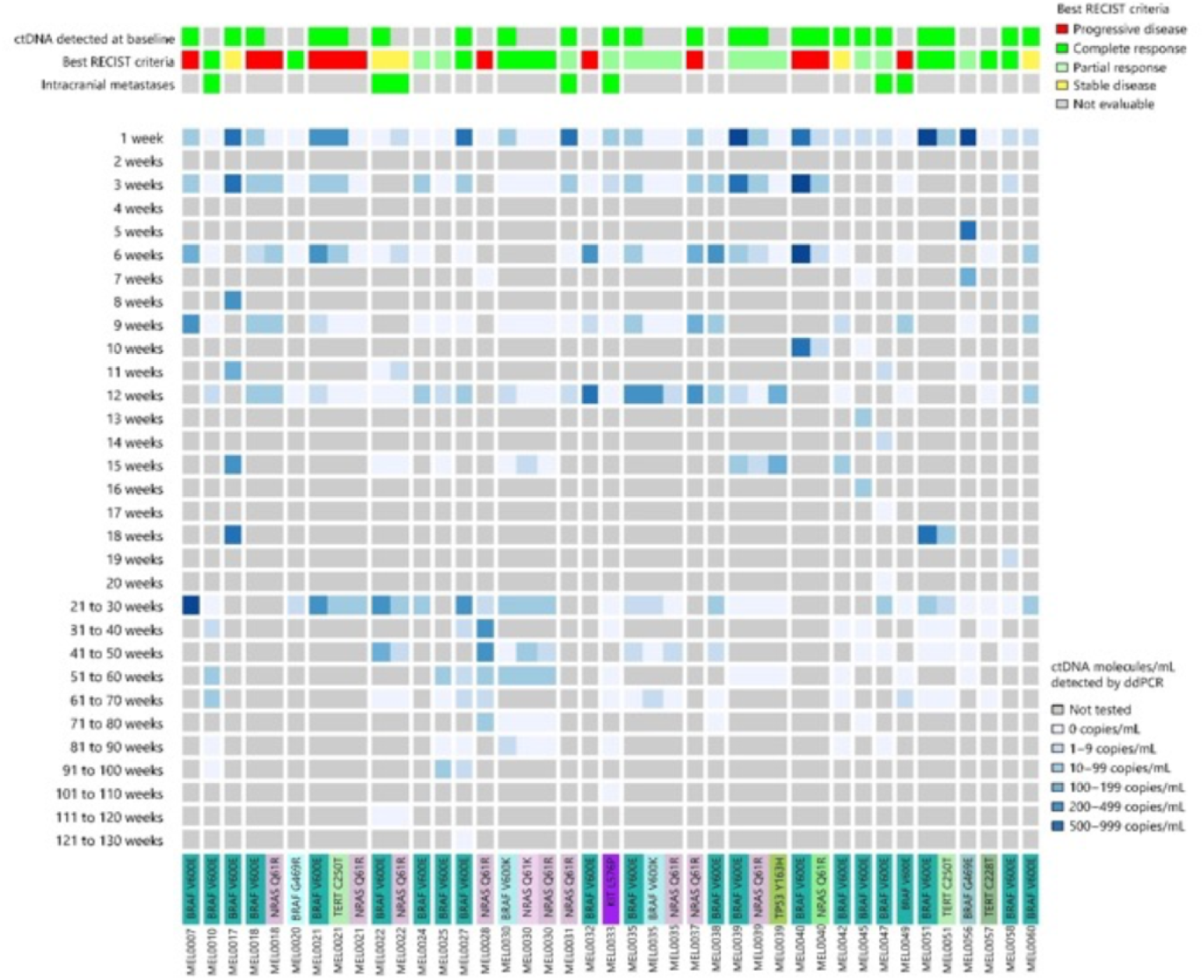
ctDNA and surveillance imaging changes over time. For each of the 29 melanoma patients, the mutations that were detected (coloured bars at bottom) and the concentration of these in each plasma sample (grey-blue scale – see key for groups) is shown. The different gene variants are colour coded. The presence of an intracranial brain metastasis is shown as either present in green or absent in grey. Monotonic ctDNA increases and decreases were observed in patients MEL007, 17, 18, 22, 27, 31, 33, 39, 51, and 56. Patients MEL0022 and MEL0031 progressed on treatment following the development of brain metastasis. Fluctuating ctDNA concentrations were identified in patients MEL0016, 21, 24, 25, 28, 32, 35, 38, 40, 42 and 49. Fluctuating low concentration ctDNA was observed in responding patients MEL0010, 20, 30, 45, 47, 58 and 60.

**Supplementary Fig. 3:**
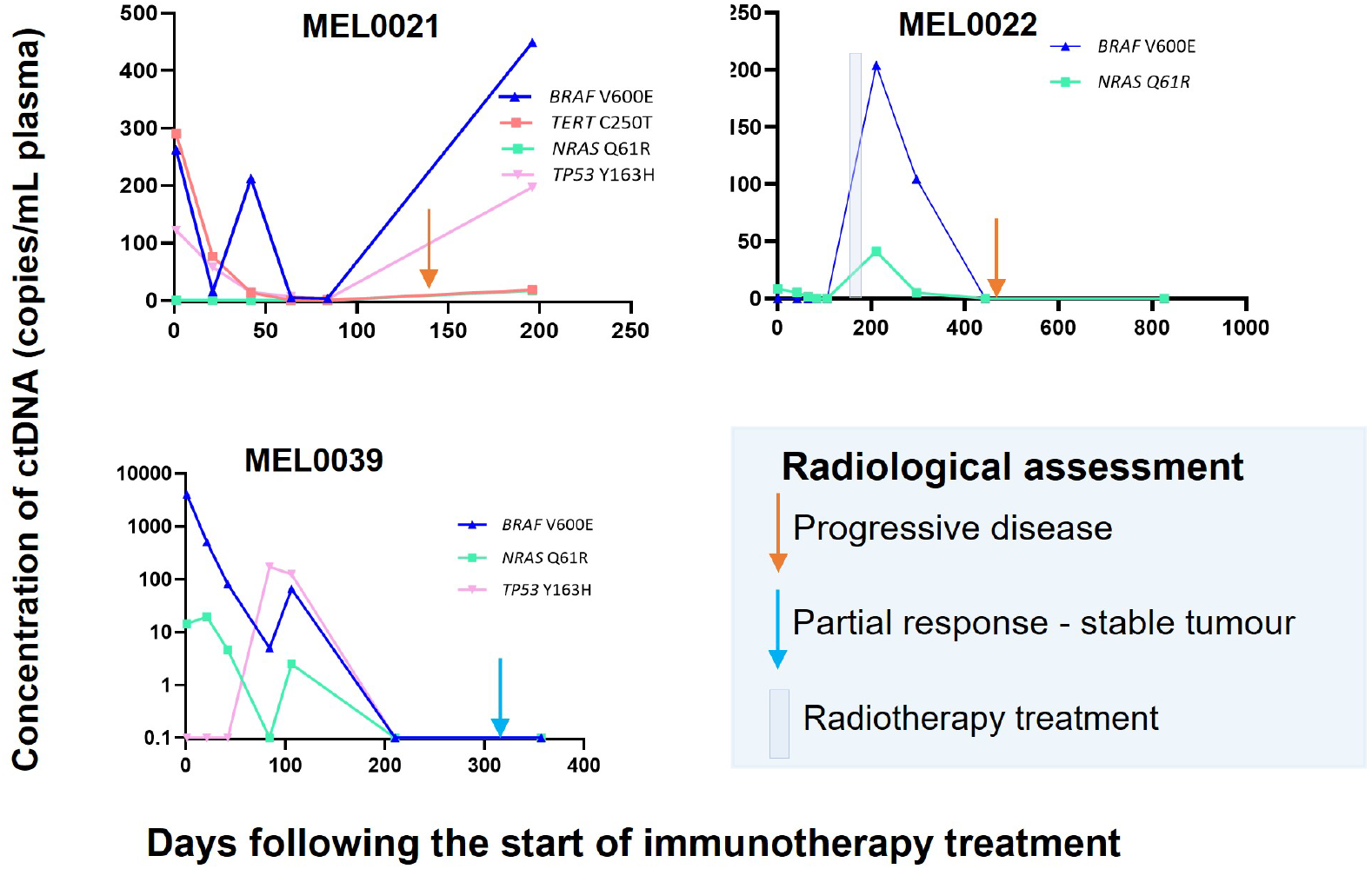
Detection of multiple mutations in the ctDNA of advanced melanoma patients. Data for three patients is shown where the longitudinal ctDNA profiles were consistent with the evolution of complex tumour heterogeneity. Patient MEL0022 demonstrated a good initial response to treatment that was reflected in the ctDNA analysis. However, this patient had subsequent intracranial disease progression. Mutations were identified by NGS conducted on plasma cfDNA samples and ddPCR assays were used to quantitate the level of ctDNA in sequential plasma samples. Concurrent *NRAS* and *BRAF* mutations were also detected for patient MEL0018 (shown in Fig. 1).

**Supplementary Fig. 4.**
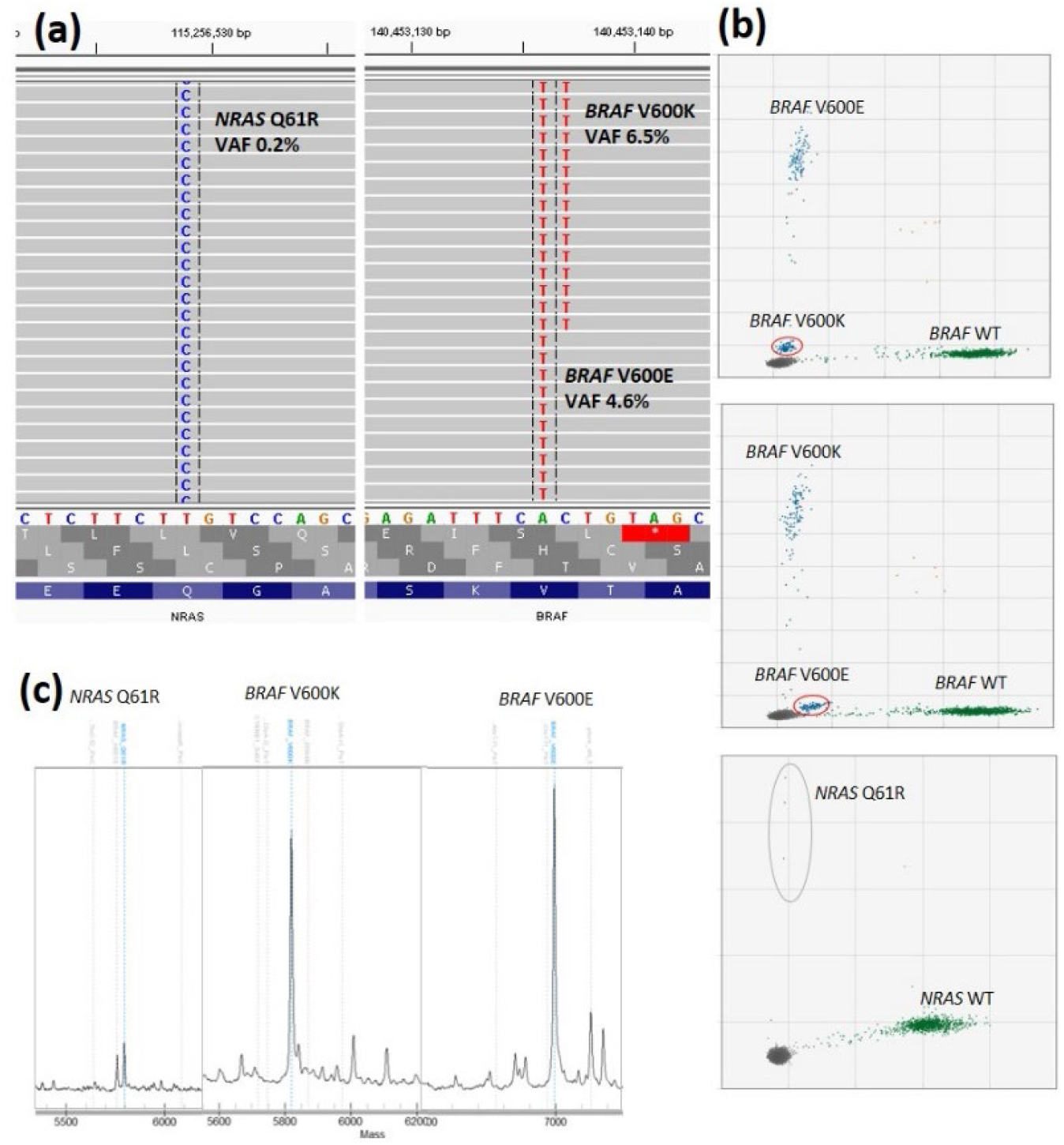
Detection of co-occurring *BRAF* and *NRAS* gene mutations in the ctDNA from a single patient. Analysis of the fifth plasma sample from patient MEL0035 is shown. (a) An Integrated Genome Browser (IGV) image of NGS sequence reads (grey bars) shows three concurrent mutations in this sample: *BRAF* V600E (c.1799A >T, 47,089 depth, 4.6% VAF), *BRAF* V600K (c.1798 AC >TT, 47,133 depth, 6.5% VAF) and *NRAS* Q61R (c.182 T >C, 25,050 depth, 0.2% VAF). The sequencing results are supported by concordant ddPCR analysis (b) and UltraSEEK MassArray results (c).

**Supplementary Fig 5.**
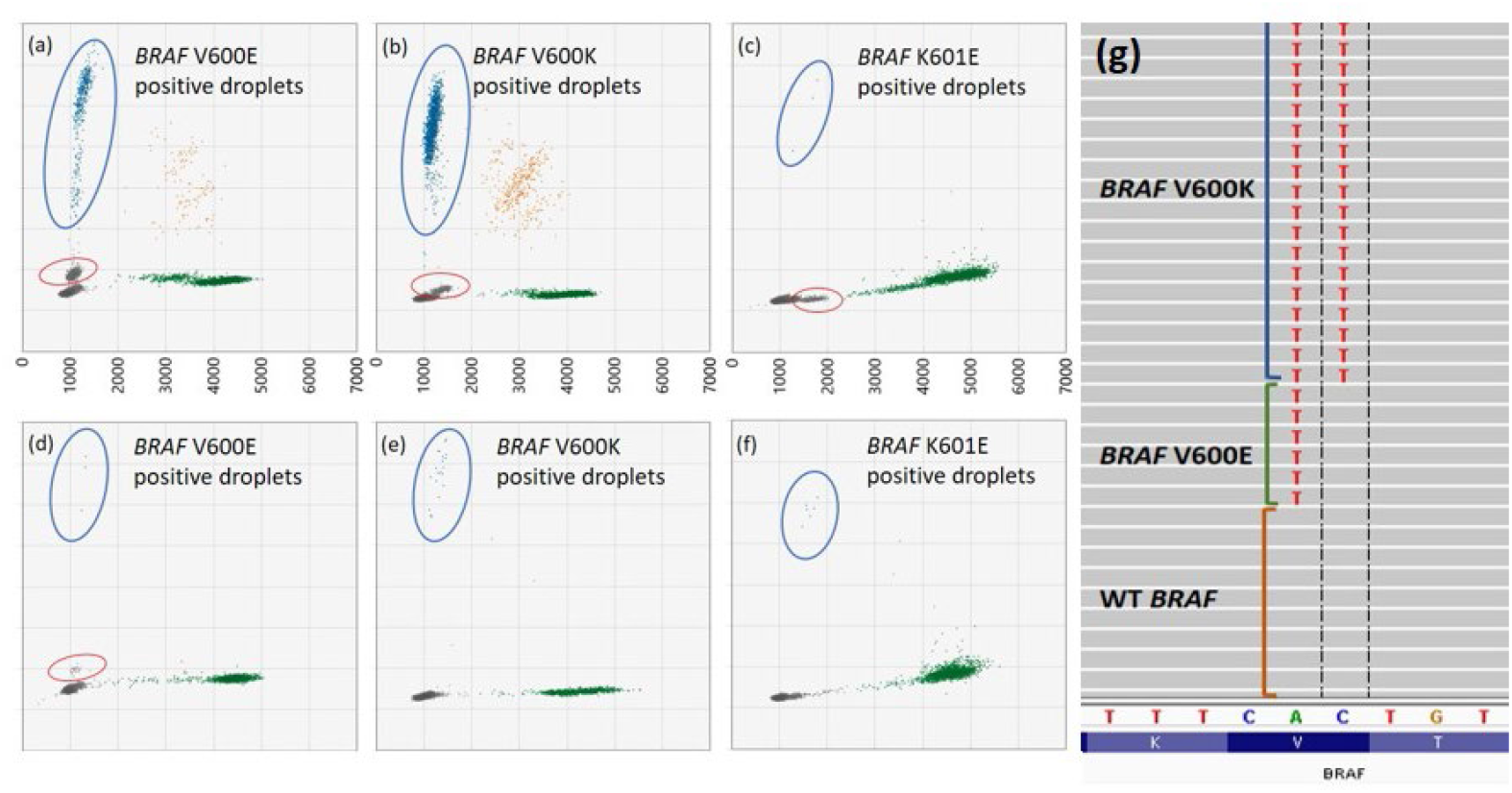
Concurrent detection of multiple *BRAF* mutations in a single patient. Analysis of tumour gDNA from patient MEL0026 with *BRAF* ddPCR assays identified (a) ctDNA for *BRAF* V600E. In addition to the main *BRAF* V600E positive droplets, a second discrete cluster (circled in red) was identified at an amplitude marginally above the negative droplets. Further analysis of tumour gDNA with ddPCR assays detected mutations for (b) *BRAF* V600K and (c) *BRAF* K601E. Similarly, when ddPCR analysis was performed on cfDNA isolated from the pre-surgical P1 plasma sample from patient MEL0026, ctDNA was identified for (d) *BRAF* V600E (e) *BRAF* V600K and (f) *BRAF* K601E. (g) IGV analysis of the tumour gDNA shows the detection of *BRAF* V600K (c.1798 AC >TT, depth 13,528, 18% VAF), *BRAF* V600E (c.1799 T>A, depth 13,538, 11% VAF) and WT *BRAF*. No *BRAF* K601E was detected by NGS analysis.

**Supplementary Fig 6.**
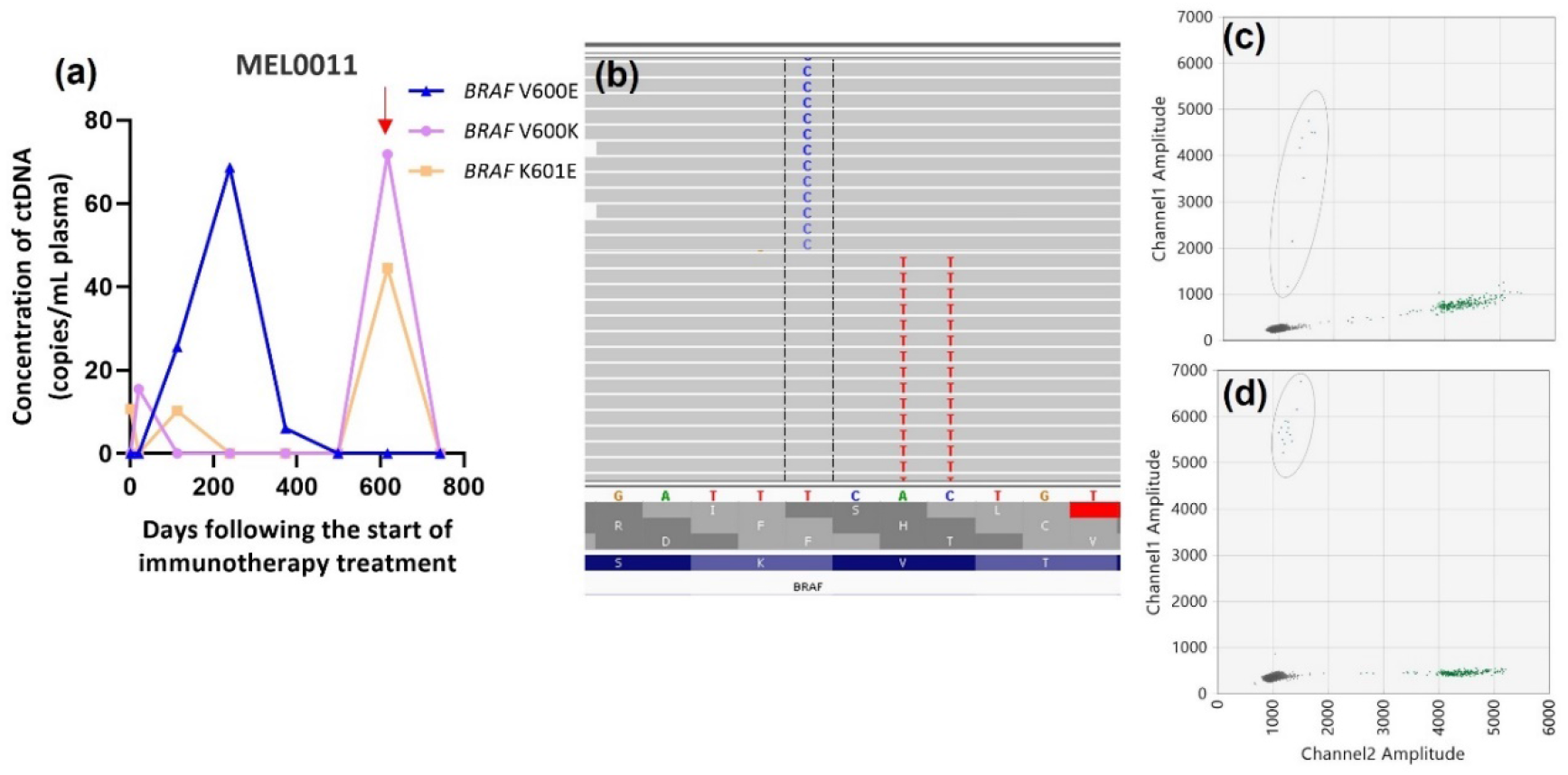
Concurrent detection of multiple *BRAF* mutations in a single patient MEL0011. (a) ctDNA analysis of MEL0011 melanoma patients showing the fluctuating concentration of three ctDNA mutations identified in the blood plasma. The red arrow shows the timing of the plasma sample analysed by NGS. (b) Sequencing performed on the cfDNA from this patient sample identified two mutations leading to a *BRAF* V600K (c.1798 AC >TT, 50,176 depth, 1.4% VAF) and *BRAF* K601E (c.1801A>G, 49,075 depth, 3.6% VAF) alteration as visualised in IGV. The presence of these mutations was confirmed by ddPCR (c) *BRAF* K601E and (d) *BRAF* V600K. In addition to the mutations identified by sequencing, a *BRAF* V600E mutation was identified by ddPCR in P3-P6 plasma cfDNA samples.

**Supplementary Fig. 7.**
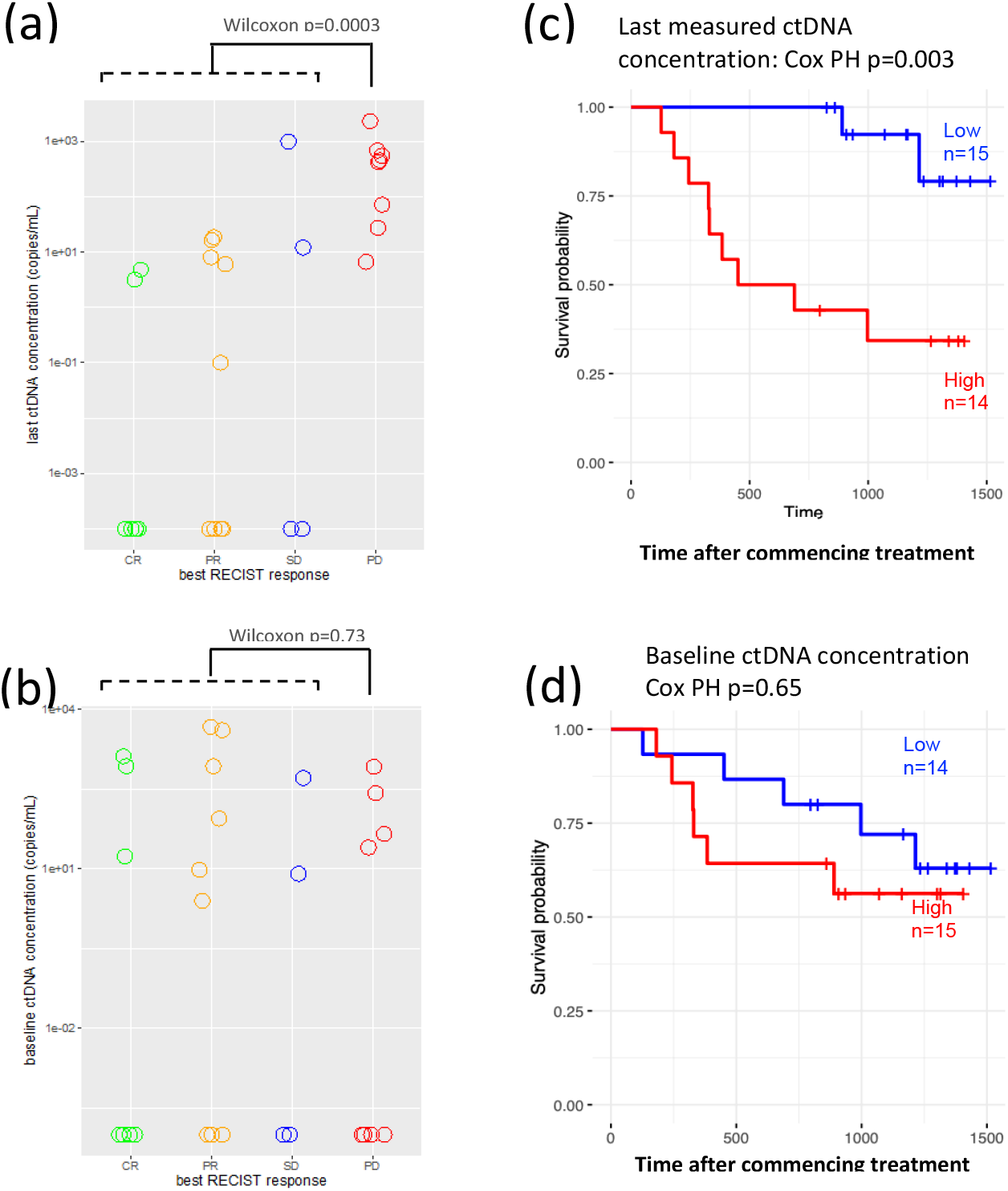
Associations between baseline or last measured ctDNA concentration and clinical outcome. The best RECIST response for each patient over the course of their immunotherapy (CR = Complete Response; PR = Partial Response; SD = Stable Disease; PD = Progressive Disease) was mapped against the final measure ctDNA concentration during the study (a and c) and baseline ctDNA concentration (b and d). Wilcoxon rank sum tests identified a significantly higher final sample ctDNA concentration in patients with progressive disease than in patients with partial or complete responses to therapy (a) but no significant differences in baseline ctDNA concentration between the four RECIST response groups (b). Cox proportional hazard models were used to assess associations between the last measured (c) or baseline (d) ctDNA concentration and patient outcome, illustrated above the Kaplan Meier graphs. A significant association was observed between final sample ctDNA concentration and patient outcome (c), but there was no significant association between baseline ctDNA concentration and patient outcome (d). Time is shown as days since the start of the first immunotherapy cycle, and the patient outcome used is overall survival. All ctDNA concentrations in this figure were determined using ddPCR. Where multiple ctDNAs were detected in a sample, the ctDNA with the highest concentration was used for this analysis.

**Supplementary Fig. 8.**
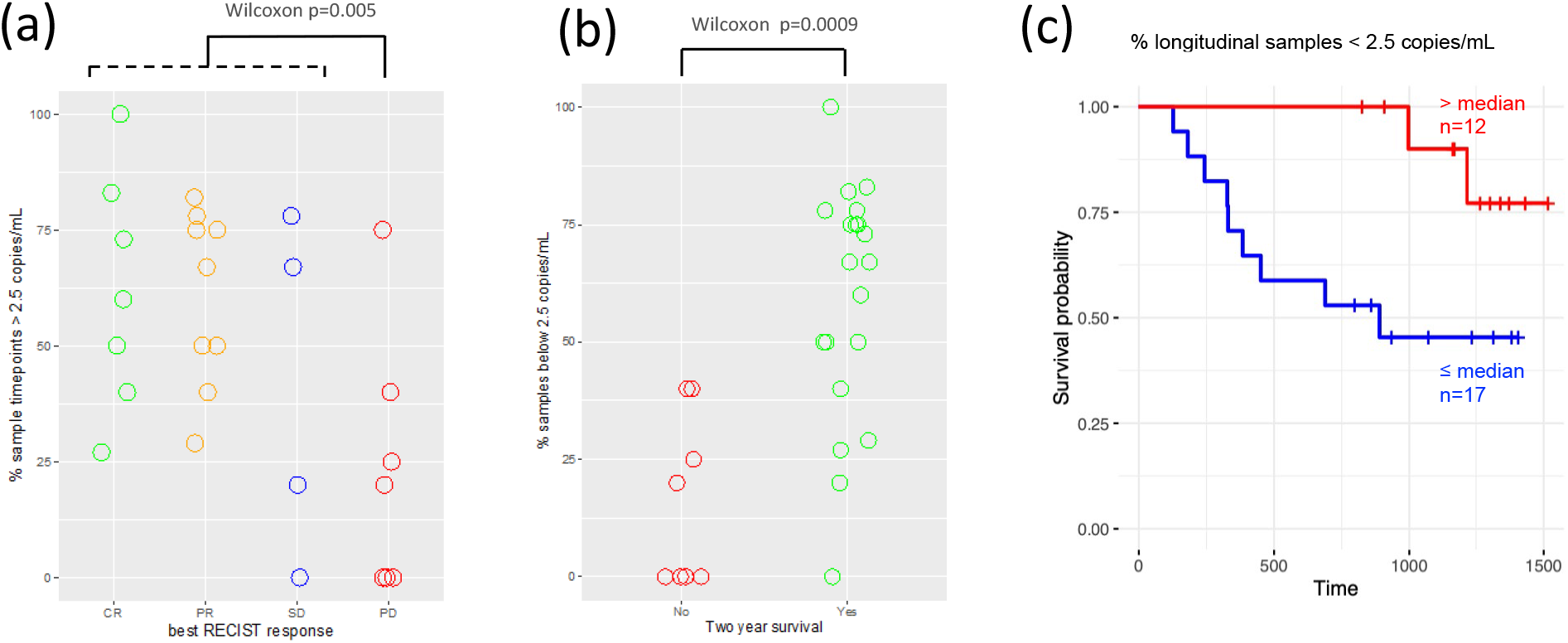
Associations between the proportion of plasma samples over the course of treatment with undetectable ctDNA and patient outcome. The best RECIST response for each patient over the course of their immunotherapy (CR = Complete Response; PR = Partial Response; SD = Stable Disease; PD = Progressive Disease) was mapped against the proportion of samples over the course of therapy with undetectable ctDNA. Wilcoxon rank sum tests identified significantly fewer samples with undetectable ctDNA in patients with progressive disease than in patients with partial or complete responses to therapy (a), and significantly fewer samples with undetectable ctDNA in patients who survival beyond 2 years (b). Cox proportional hazards analysis identified a significant positive association between patient outcome (overall survival) and the proportion of plasma samples over the course of therapy with undetectable ctDNA, shown above the Kaplan Meier graphs in which patients are grouped above (high - red) and below (low – blue) the median (c). All ctDNA concentrations in this figure were determined using ddPCR and the limit of detection for ctDNA was set at 2.5 copies/mL. Where multiple ctDNAs were detected in a sample, the ctDNA with the highest concentration was used for this analysis.

**Supplementary Fig. 9.**
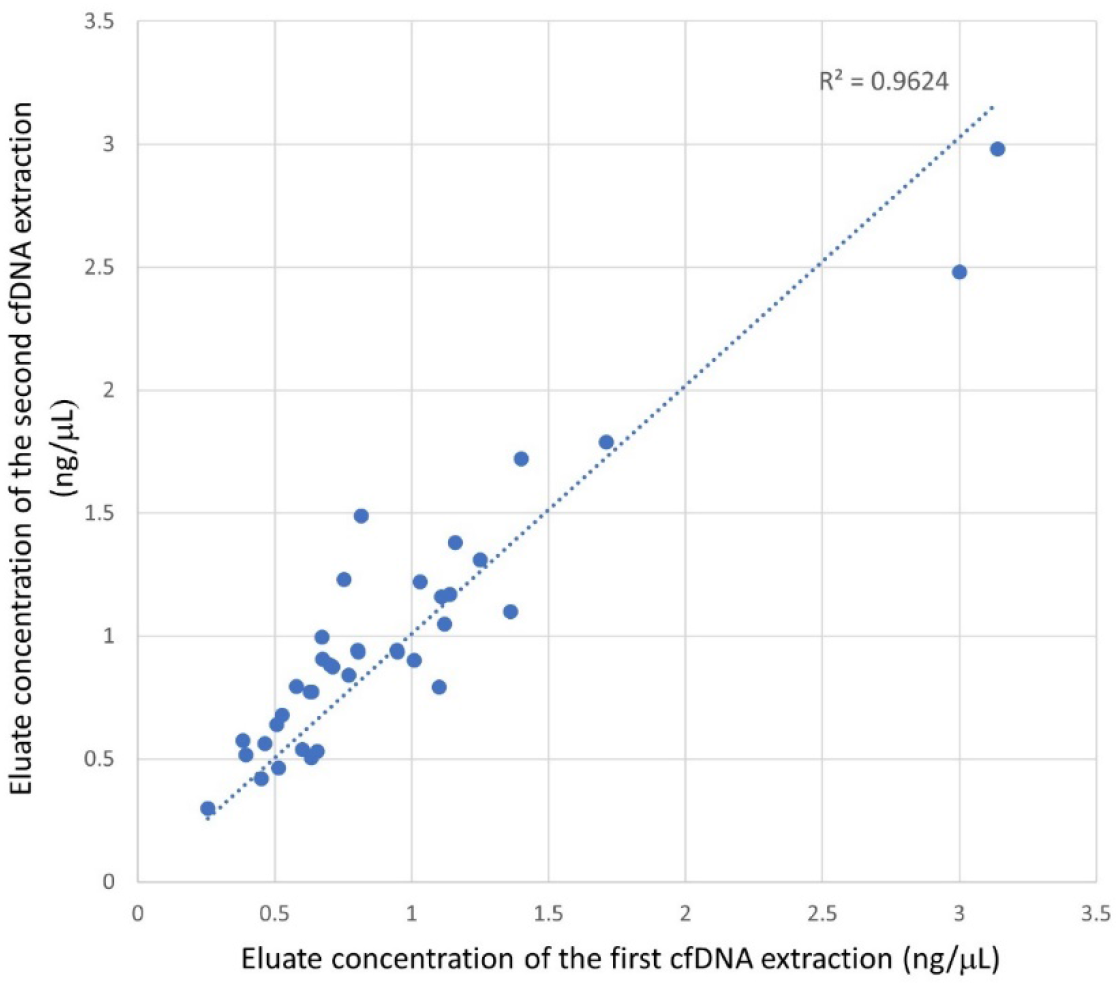
Scatterplot showing the concentration of cfDNA following extraction of two aliquots of patient plasma on separate days. cfDNA was isolated from 38 patient plasma samples. The variability in the eluate cfDNA concentration determined by Qubit fluorometry is plotted as ng/μL. The correlation coefficient was R^2^=0.9624. The trendline is shown in blue.

**Supplementary Fig. 10:**
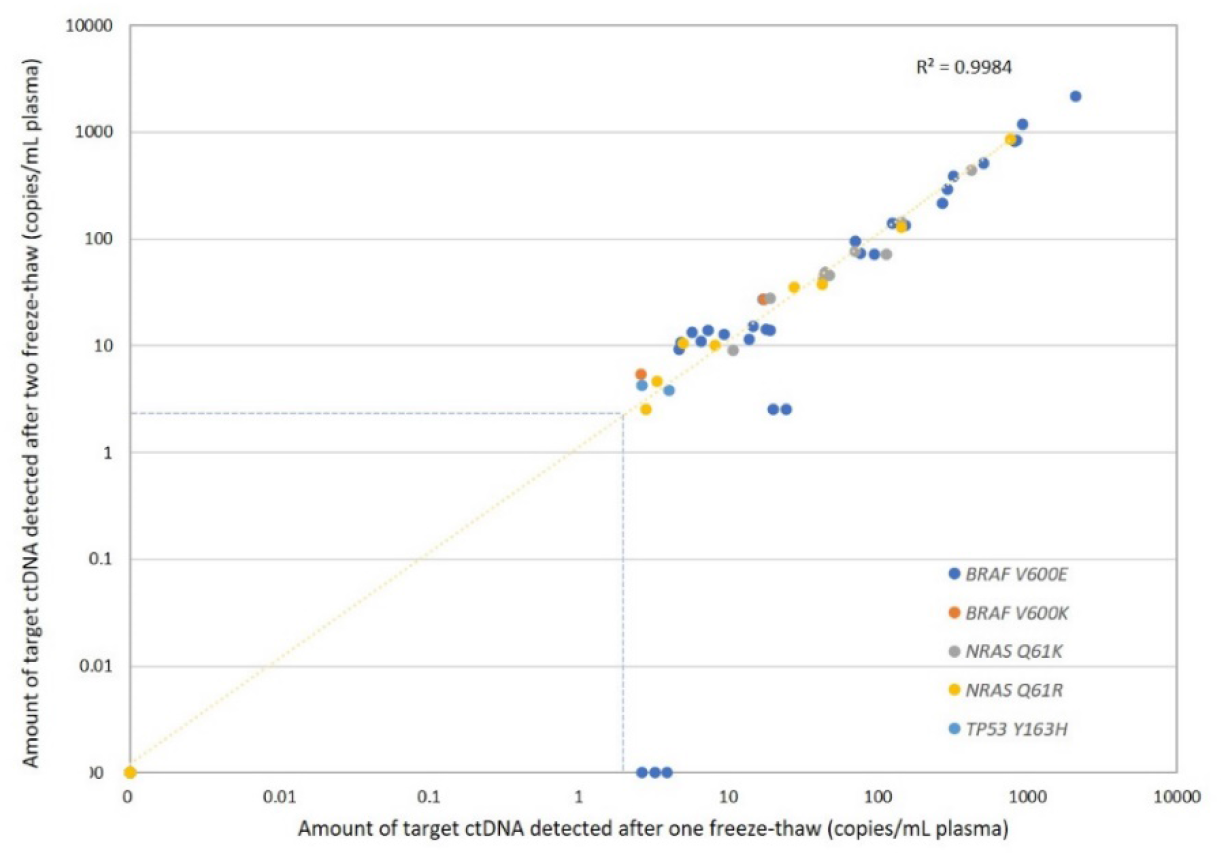
Scatterplot showing the quantitation of ctDNA following one or two freeze-thaw cycles. The concentration of ctDNA by ddPCR assay following one freeze-thaw is plotted on the x-axis. The concentration of ctDNA in the same sample by ddPCR assay following a second freeze-thaw is plotted on the y-axis. Each coloured circle represents a different mutation quantitated by ddPCR assay. The correlation coefficient was R^2^ =0.9984. The blue dotted line that is drawn represents the threshold for reporting a mutation of >2.5 copies/mL input plasma. The trendline is shown in yellow.

**Supplementary Fig. 11.**
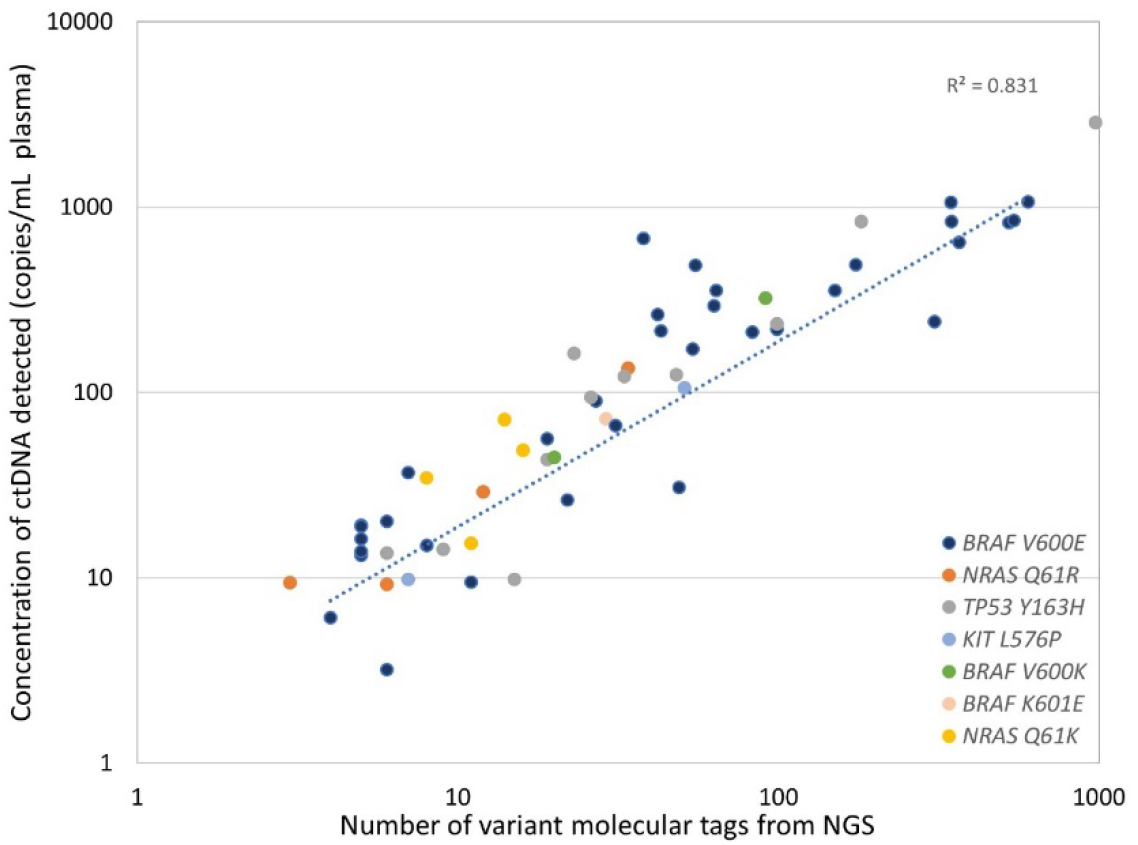
Scatterplot showing the correlation between the number of VMTs detected by NGS and the quantitation of ctDNA by ddPCR assay. Following NGS of patient cfDNA samples, the number of VMTs reported for a mutation is plotted on the x-axis. For all mutations detected by NGS, the corresponding ddPCR assay was used to quantitate this mutation in the patient cfDNA and is presented on the y-axis as copies/mL plasma. The coloured circles represent the different mutations identified by NGS and quantitated with ddPCR assays. The correlation coefficient was R^2^ =0.831. The trendline is shown in blue.

**Supplementary Fig. 12.**
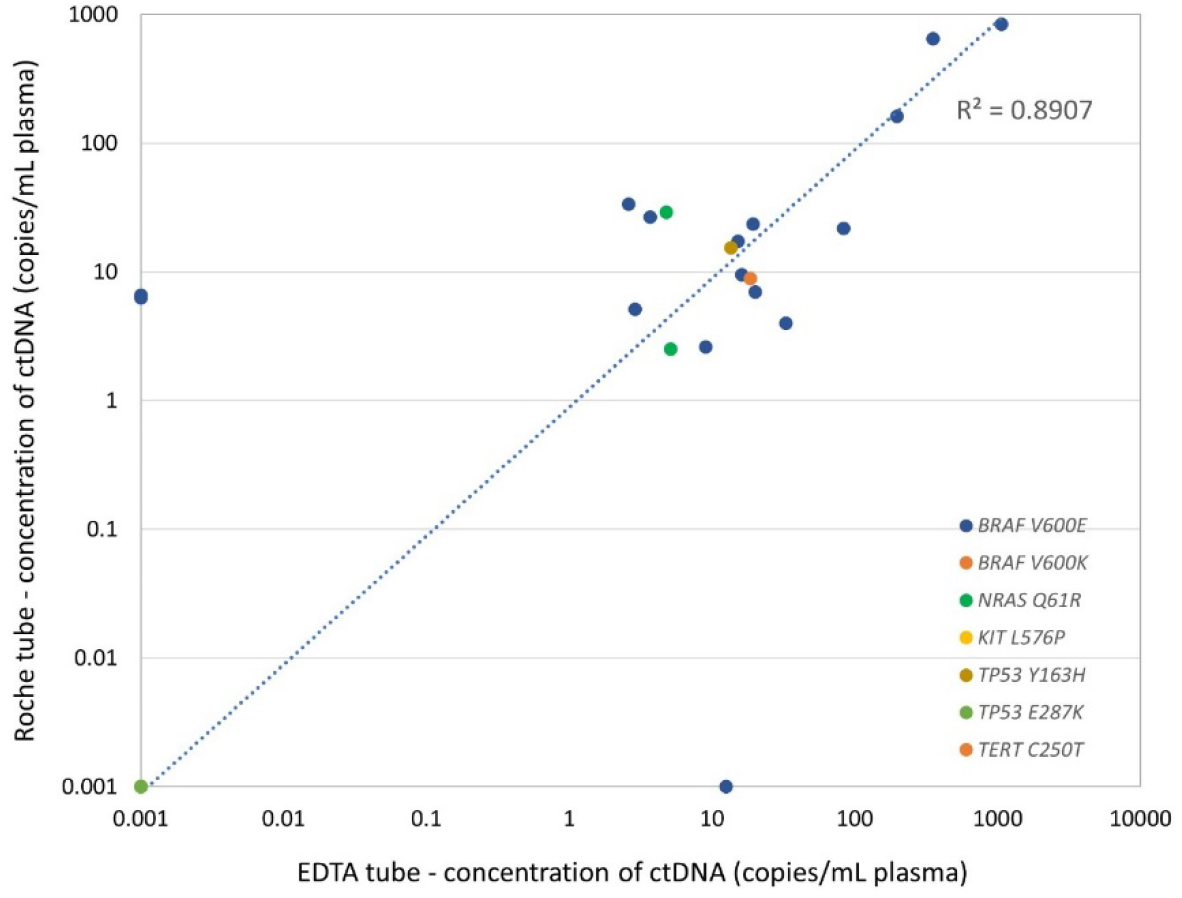
Correlation between ctDNA quantitated in EDTA tubes vs Roche Cell-free collection tubes processed after seven days. The quantitation by ddPCR assay for mutation detection in EDTA collected blood samples is plotted on the x-axis. The Roche tube collected blood samples are plotted on the y-axis. For each patient plasma sample, the known mutation was quantitated by ddPCR and reported as copies/mL plasma. The coloured circles represent the different mutations identified and quantitated by ddPCR assay. The correlation coefficient was R^2^=0.8907. The trendline is shown in blue.

